# Distinguishing Age-specific Patterns in Comorbidities of Obstructive Sleep Apnea Using Real-World Data

**DOI:** 10.64898/2026.05.20.26352336

**Authors:** Matthew O. Goodman, Raichel M. Alex, Scott A. Sands, Ali Azarbarzin, Salma Batool-Anwar, Milena K. Pavlova, Lawrence J. Epstein, Susan Redline, Brian E. Cade

## Abstract

Obstructive sleep apnea (OSA) is associated with a wide range of comorbidities, but the extent to which these follow predictable, age-dependent patterns is not well understood. Identifying such patterns could provide insight into OSA heterogeneity and its links to physiological measures of OSA. We trained age-dependent topic models (ATM) on longitudinal electronic health records from 36,426 patients with OSA in the Mass General Brigham Biobank. ATM organizes incident diagnoses into distinct comorbidity “topics,” whose age-specific disease loadings represent predictive patterns linking related diagnoses across the life course. We applied the trained model to compute individual-level topic scores in independent data: a cohort of 11,689 OSA cases and 22,695 matched controls, and a cohort of 6,220 patients with polysomnography (PSG)-derived physiological measures. We identified 19 distinct age-dependent comorbidity profiles, all significantly associated with OSA case status (FDR-adjusted p<0.05). Topics reflected recognizable clusters including metabolic, neuropsychiatric, and immune-mediated conditions, and several were distinguished by age-of-onset of key comorbidities, such as early- vs late-onset asthma. Seventeen of the 19 topics were significantly associated with at least one of 13 PSG-derived physiological measures, including associations between cardiometabolic topics and the apnea-hypopnea index, sleep apnea specific hypoxic burden, and respiratory event-specific heart rate burden. These findings indicate that age-dependent comorbidity patterns distinguish meaningful OSA subtypes with differing prognoses and endophenotype associations. ATM offers insight into complex OSA comorbidity and suggests that age-informed, topic-based stratification may improve individualized risk assessment, interpretation of PSG findings, and targeting of clinical interventions.

**One Sentence Summary:** Analysis of age-specific patterns in comorbidities of obstructive sleep apnea reveals insights for personalized care and risk stratification.

## INTRODUCTION

Obstructive sleep apnea (OSA) is a common and serious sleep disorder characterized by repetitive upper airway obstruction during sleep, resulting in intermittent hypoxemia and sleep fragmentation (*1*). OSA is associated with a substantial burden of comorbid disease across cardiometabolic, cardiovascular, respiratory, and neuropsychiatric domains, including conditions such as hypertension, type 2 diabetes, coronary artery disease, cerebrovascular disease, and cognitive decline (*2*). The heterogeneous nature of OSA, evident in its clinical presentation, comorbidity profiles, and long-term outcomes, creates challenges for risk stratification and clinical management (*3*). Recent therapeutic advances, including pharmaceutical agents and neurostimulation devices, have expanded opportunities for personalized treatment, emphasizing the need to develop strategies that match patients to the most appropriate intervention (*4–6*). Recent work to advance understanding of OSA heterogeneity has investigated variation in presentation, pathogenesis and adverse outcomes and how these evolve across the adult age span (*7–9*). In this context, study of distinct age-related patterns in comorbidities may highlight differences in OSA useful for risk stratification. Age-dependent topic modeling (ATM), described below, offers a promising framework to characterize such patterns.

OSA has been linked to a diverse set of comorbid conditions through various studied pathways, including intermittent hypoxia, autonomic dysregulation, and systemic inflammation (*1, 10*). Given OSA’s broad systemic effects, it is possible that research has overlooked important associated conditions, as suggested by recent phenome-wide association studies (PheWAS) (*11*). Nonetheless, prior OSA multimorbidity research has faced two key limitations. Prospective cohort studies of incident events have considered a limited subset of well-recognized associated conditions, e.g. obesity, hypertension, diabetes, heart disease, stroke, depression, and cognitive decline. Meanwhile, phenome-wide analyses of electronic health records (EHR) reveal associations with hundreds of additional diagnoses, yet these studies have been limited by either examining each condition independently (*11*) or by deploying cross-sectional clustering methods (*12*). As such, these approaches may have overlooked temporal sequences and age-specific patterns in comorbidity development that could be informative for risk stratification.

Systematic reviews of multimorbidity research emphasize the predictive value of longitudinal analyses linking co-occurring diseases to one another and to long-term outcomes (*13, 14*). These trajectory-based approaches in various chronic disease contexts have identified clinically meaningful patient groups with distinct comorbidity clusters and temporal sequencing. For example, trajectories with cardiometabolic multimorbidity preceding later psychiatric and neurodegenerative disorders (15) can be distinguished from trajectories beginning with depression and proceeding to cardiovascular disease (*13, 15*). In OSA, prior phenotyping efforts have applied cluster analysis to symptom profiles, PSG metrics, biomarker panels or comorbidities (*16–22, 12*). But these approaches have been largely cross-sectional, based on features recorded at a single time point (*23, 24, 12*). Use of EHR-derived comorbidity patterns can provide a window into longitudinal patient health while repurposing existing clinical data, in some cases hundreds of conditions across decades of records.

Age-dependent topic modeling (ATM) is a Bayesian method that infers age-dependent patterns from incident diagnoses in longitudinal EHR data (*25*). ATM provides a method that can organize co-occurring conditions into longitudinal trajectories, or topics, accounting for age-of-onset, as well as predict risk of future incident comorbidities within the same topic. Specifically, the method trains several linear and non-linear models with varying parameters, selecting a final model based on measured performance predicting incident diagnoses in validation data (Methods). These topics link co-occurring diagnoses to generate predictive age-dependent patterns over the life course. For example, a hypothetical neuropsychiatric topic might progress from mood disorders in early adulthood to insomnia in middle age and cognitive decline in later life. ATM topic scores indicate individuals’ risk of developing further incident diagnoses within that topic. These topic scores can be calculated using the learned age-specific disease weights for each topic, in combination with an individual’s longitudinal EHR, and used to predict incident diagnoses at future ages. Importantly, ATM-derived topics suggest stable, trait-like patterns that could serve as biomarkers for personalized intervention strategies targeting specific classes of later life outcome. Applied to OSA, ATM provides a framework to characterize age-related comorbidity trajectory patterns and assess their associations with disease status and pathophysiology.

Previous application of ATM demonstrated utility for identifying predictive age-specific patterns of disease incidence across the life course, including an application of ATM to UK Biobank and All of Us cohorts, finding that topics were predictive, transportable, and associated with distinct genetic subtypes (*25*). Disease subtypes defined by topic membership, distinguishing both comorbidity groupings and age-of-onset presentations, exhibited distinct genetic architectures, heritability patterns, and polygenic risk scores (*25*).

In this work, we apply ATM to identify patterns in life-course comorbidity progression in OSA cases, using EHR records from the Mass General Brigham health system. In an independent data set, we then test associations between these topics and OSA case-control status as well as selected polysomnography (PSG) measures of OSA (*26*). These analyses provide a unique framework for characterizing age-dependent patterns in comorbidities of OSA that may prove useful for clinical risk stratification and precision medicine. We hypothesize that ATM topics, trained in OSA cases on OSA-associated comorbidities, i) reveal both recognized and novel predictive age-dependent comorbidity patterns that ii) differentially associate with OSA status in independent testing data, with varying effect sizes suggesting groups of comorbidities where risk and/or age of onset most differs by OSA status, and iii) differentially associate with specific OSA endophenotypes, with differing association profiles suggesting potentially differing underlying mechanisms and targets for risk stratification.

## RESULTS

### Study Design and Population

We analyzed data from 163,220 total patients in the MGB system, merging clinical research data, and electronic health records (EHR) (*27*). All patients met inclusion criteria including a minimum of three distinct encounter dates (Methods). Clinical ICD diagnosis codes were obtained from EHR, to which we applied the ‘phecode’ clinical ontology (*28*) to group related codes. We then used the Multimodal Automated Phenotyping (MAP) EHR phenotyping algorithm to improve the diagnostic accuracy of comorbidities (*29*) (Methods).

Exposures consisted of OSA status or PSG measures. OSA status was ascertained using PheVis (*30*) in a validated algorithm incorporating EHR codes and clinical notes as well as available AHI values ≥ 15. OSA cases younger than 18 years old at the time of diagnosis (first date of PSG or EHR diagnosis) were excluded due to paucity of data and lack of follow-up into adulthood. Controls were matched 2:1 to cases by matching on BMI and age within specified tolerances, and propensity scores for remaining covariates (*31*), ensuring overlapping covariate distributions (Methods). This resulted in 48,251 ascertained OSA cases and 96,502 controls. Additionally, to reduce confounding, all models were adjusted for the matching covariates. Adjustment and matching covariates consisted of age (at PSG or at OSA diagnosis), sex, BMI, and self-reported ancestry as well as measures of healthcare utilization (age at first EHR encounter and number of total encounters).

To protect validity of association testing and preserve Type-1 error, we defined fixed non-overlapping training and testing cohorts for ATM analysis (Methods). Specifically, ATM training and model-validation data consisted of 36,426 ascertained OSA cases (case-only analysis). Testing data consisted of two groups: first, an OSA case-control testing dataset consisting of 11,689 OSA ascertained cases and 22,695 matched controls, having complete covariate data; second, an additional PSG testing dataset of 6,220 individuals, consisting of patients with complete covariate data and available clinical PSG from the Brigham and Women’s Faulkner Hospital sleep clinic, selected without regard to OSA status (3,594 with AHI ≥15 and 2,626 with AHI <15). In the PSG dataset we tested associations between ATM topics and 13 selected OSA-related continuous PSG measures (*26*) (Methods).

Distributions of demographic and health-related covariates by sub-cohort (Table 1) confirm success of the matching design (Methods) and align with expectation for distribution in OSA cases, albeit with some depletion of unmatched high-BMI cases. The Faulkner PSG cohort had fewer males (38.1% vs 54.8% among cases and controls) and differing distribution of ancestries, but similar age, BMI, and healthcare encounters per year.

**Fig. 1.**
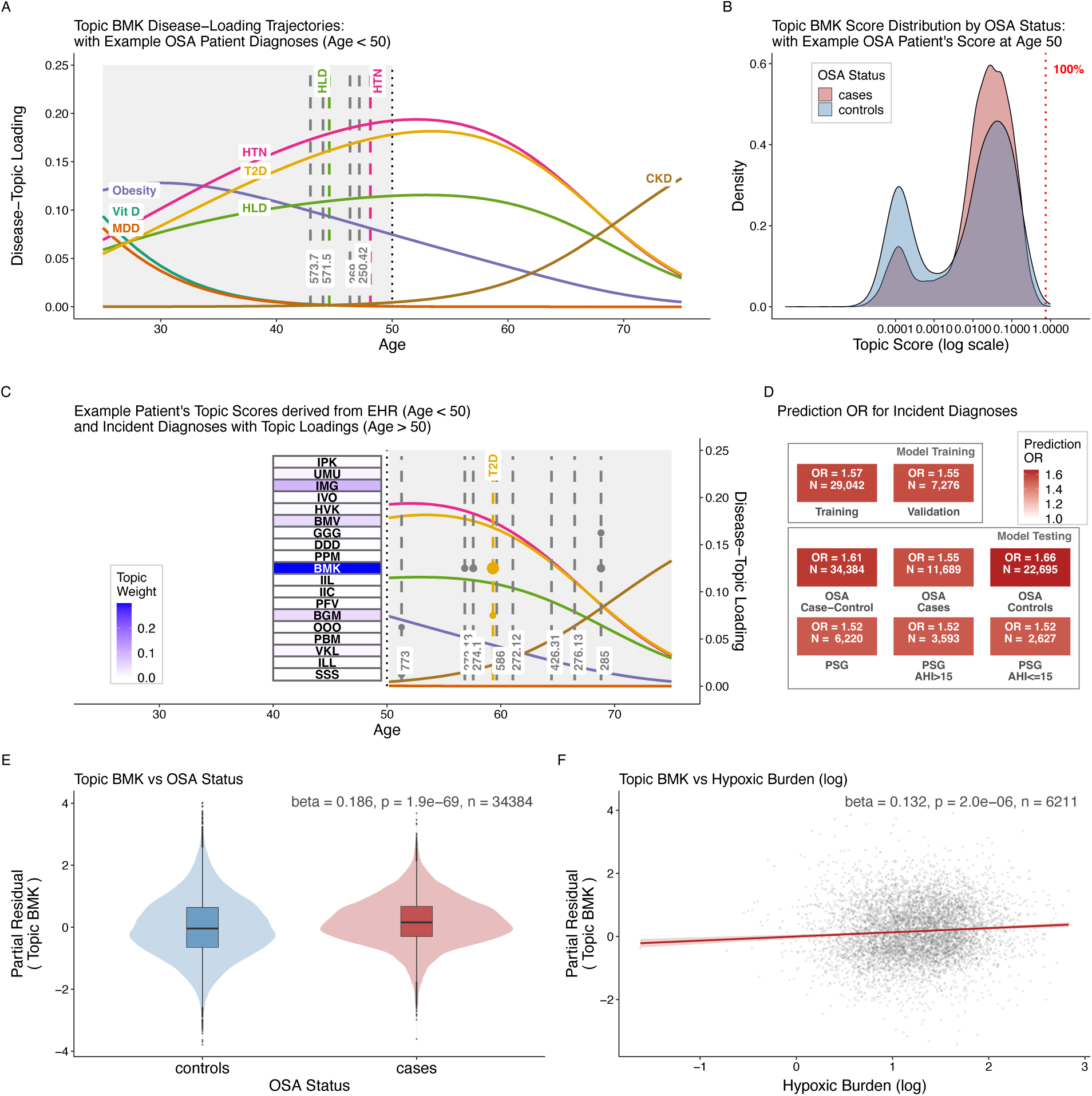
Key figure: ATM Comorbidity Modeling: Example Topic Profile (BMK), Prediction, and Association Analyses. **(A)** Disease-topic loading trajectories for seven diseases associated with Topic BMK (Vit D deficiency, MDD, Obesity, HLD, T2D, HTN, CKD), showing how each disease’s contribution to the topic varies across age. Dashed vertical lines indicate the age at incident diagnosis for a synthetic example OSA patient with high BMK topic score (0.3), simulated from the learned ATM for illustration. **(B)** Topic BMK score distribution among OSA cases and controls, displayed on a log scale. The red dotted line marks the example patient’s BMK topic score at age 55, with the corresponding percentile among cases shown. **(C)** This panel shows the example patient’s inferred ATM topic scores (left) alongside post-cutoff incident diagnoses (right). The blue heatmap (left) displays the patient’s topic score for each topic, computed from their EHR history up to age 55, using the learned ATM with weights representing 305 EHR diagnoses, across 19 topics, from ages 25 through 90. The heatmap displays the inferred weight of each of the 19 ATM topics based on pre-cutoff diagnoses, with topic abbreviations labeled within each cell. In the post-cutoff region (right), dashed vertical lines indicate incident diagnoses, with colored dots at each topic row representing their importance (disease-topic loading) at the given age. Loading curves from Panel A are shown again for context. **(D)** Population-level prediction odds ratios (OR, top 10% threshold) across eight study sub-cohorts, organized by model training and model testing groups. Each tile displays the prediction OR (color intensity) and sample size. The prediction OR quantifies how well ATM-predicted disease rankings identify a patient’s next diagnosis compared to a prevalence-based baseline prediction (See Methods). **(E)** Covariate-adjusted partial residual plot of Topic BMK score versus OSA status (cases vs. controls), adjusting for demographic and clinical covariates (see Methods). **(F)** Covariate-adjusted partial residual plot of Topic BMK score versus log-transformed hypoxic burden in the Faulkner PSG cohort, with the same covariate adjustment as Panel E. **Abbreviations:** *Clinical:* OSA, Obstructive Sleep Apnea; PSG, polysomnography; *Diagnoses:* CKD, chronic kidney disease; HLD, hyperlipidemia; HTN, hypertension; MDD, major depressive disorder; T2D, type 2 diabetes; Vit D, vitamin D deficiency. *Model:* ATM, Age-dependent Topic Modeling; BMK, BMI-Metabolic-Kidney-Cardiovascular topic.

**Table 1a.** Distributions in MGB OSA Case-Control Testing Cohort. Demographic characteristics, covariates, and ATM topic risk-scores for OSA cases and matched controls in the held-out MGB testing cohort. Cases were matched 2:1 to controls using age, sex, BMI, self-reported ancestry, and healthcare utilization (age at first encounter, total encounters). Topic risk-scores (T01–T19) represent individualized scores for each ATM-derived comorbidity trajectory, computed from an individual’s longitudinal EHR. Scores quantify how closely an individual’s pattern of incident diagnoses, including age at each diagnosis, matches the age-dependent progression characteristic of each topic; higher scores indicate greater concordance with the topic’s trajectory and predict elevated risk of future incident diagnoses within that topic. Continuous variables are presented as median [IQR] or mean (SD). Population represents self-reported ancestry group. Three-letter topic abbreviations denote characteristic comorbidity patterns (see Table 2a for topic descriptions). Raw topic scores are presented here, but topics were rank normalized (RN) to standardize prior to association testing.

| Variable |  | Cases |  | Controls |  |
| --- | --- | --- | --- | --- | --- |
| n |  | 11689 |  | 22695 |  |
| Age at first encounter | (median [IQR]) | 46.2 | [35.6, 56.3] | 48.2 | [37.1, 57.9] |
| N encounters | (median [IQR]) | 130.0 | [63.0, 222.0] | 84.0 | [30.0, 192.0] |
| Age | (median [IQR]) | 57.6 | [47.2, 67.0] | 57.8 | [47.4, 67.1] |
| BMI | (median [IQR]) | 29.9 | [26.7, 34.1] | 30.9 | [27.3, 35.2] |
| Population (%) |  |  |  |  |  |
| African |  | 1069 | ( 9.1) | 1446 | ( 6.4) |
| Asian |  | 337 | ( 2.9) | 560 | ( 2.5) |
| European |  | 9085 | (77.7) | 18776 | (82.7) |
| Hispanic |  | 392 | ( 3.4) | 493 | ( 2.2) |
| Other |  | 806 | ( 6.9) | 1420 | ( 6.3) |
| Sex (male) | (mean (SD)) | 0.548 | (0.498) | 0.547 | (0.498) |
| T01 IPK | (mean (SD)) | 0.047 | (0.067) | 0.074 | (0.127) |
| T02 UMU | (mean (SD)) | 0.052 | (0.077) | 0.051 | (0.098) |
| T03 IMG | (mean (SD)) | 0.058 | (0.071) | 0.046 | (0.072) |
| T04 IVO | (mean (SD)) | 0.055 | (0.072) | 0.054 | (0.097) |
| T05 HVK | (mean (SD)) | 0.054 | (0.096) | 0.048 | (0.106) |
| T06 BMV | (mean (SD)) | 0.046 | (0.070) | 0.049 | (0.093) |
| T07 GGG | (mean (SD)) | 0.048 | (0.066) | 0.046 | (0.082) |
| T08 DDD | (mean (SD)) | 0.074 | (0.119) | 0.056 | (0.128) |
| T09 PPM | (mean (SD)) | 0.058 | (0.090) | 0.043 | (0.098) |
| T10 BMK | (mean (SD)) | 0.047 | (0.063) | 0.047 | (0.082) |
| T11 IIL | (mean (SD)) | 0.041 | (0.069) | 0.033 | (0.075) |
| T12 IIC | (mean (SD)) | 0.058 | (0.071) | 0.056 | (0.094) |
| T13 PFV | (mean (SD)) | 0.059 | (0.086) | 0.052 | (0.097) |
| T14 BGM | (mean (SD)) | 0.048 | (0.075) | 0.046 | (0.089) |
| T15 OOO | (mean (SD)) | 0.061 | (0.089) | 0.078 | (0.147) |
| T16 PBM | (mean (SD)) | 0.048 | (0.065) | 0.041 | (0.080) |
| T17 VKL | (mean (SD)) | 0.047 | (0.085) | 0.068 | (0.122) |
| T18 ILL | (mean (SD)) | 0.045 | (0.067) | 0.050 | (0.089) |
| T19 SSS | (mean (SD)) | 0.053 | (0.086) | 0.061 | (0.125) |

**Table 1b.**
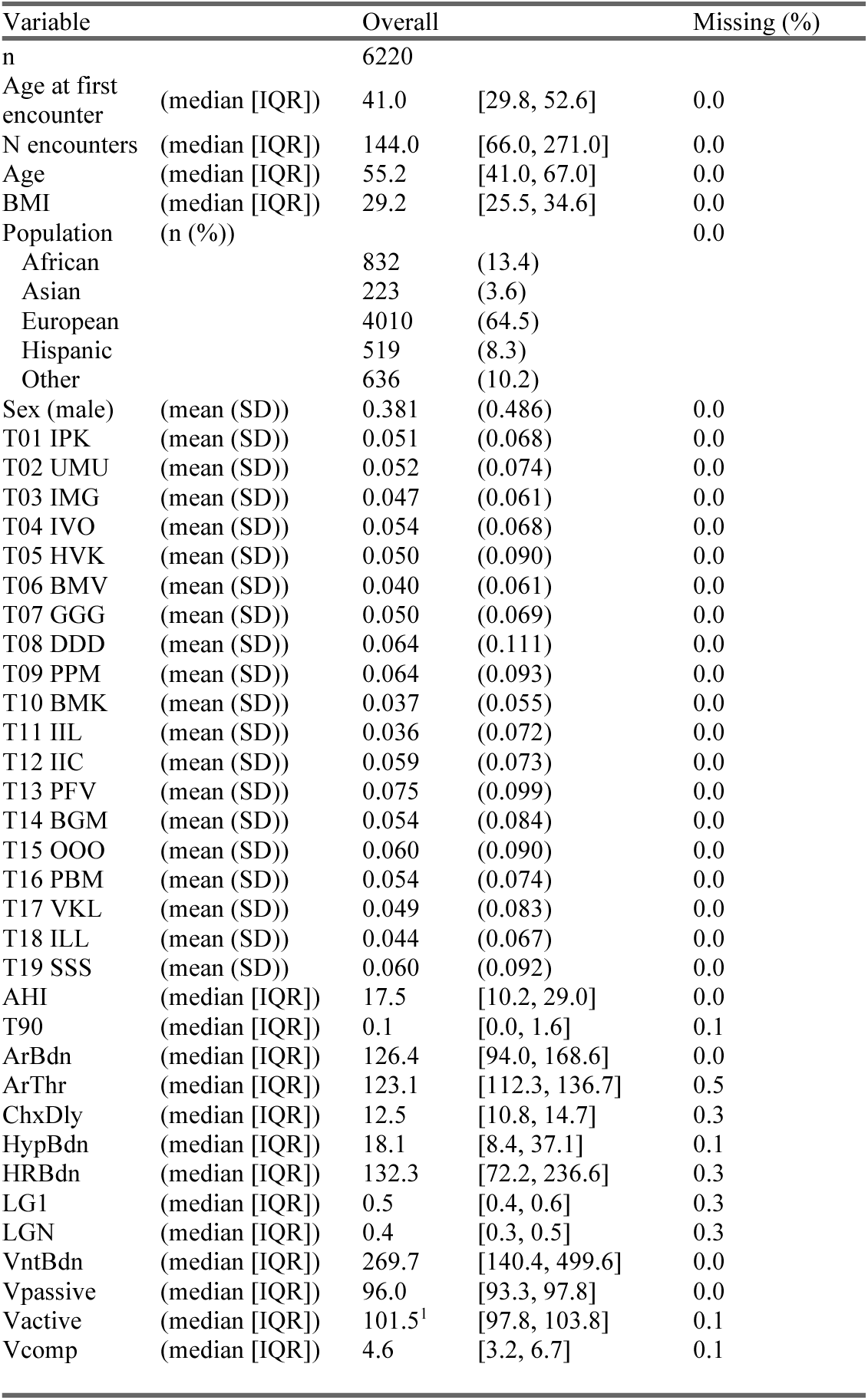
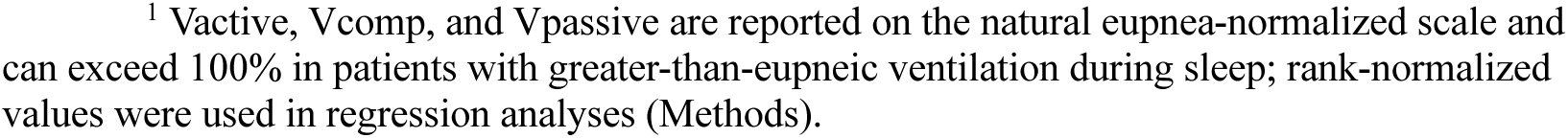
Distributions in the Polysomnographic Testing Cohort. Demographic characteristics, covariates, ATM topic risk-scores, and PSG measures for individuals with sleep studies in the held-out PSG testing cohort. (See Table 2a for ATM topic descriptions; see Table 4 for PSG measure descriptions).

| Variable |  | Overall |  | Missing (%) |
| --- | --- | --- | --- | --- |
| n |  | 6220 |  |  |
| Age at first encounter | (median [IQR]) | 41.0 | [29.8, 52.6] | 0.0 |
| N encounters | (median [IQR]) | 144.0 | [66.0, 271.0] | 0.0 |
| Age | (median [IQR]) | 55.2 | [41.0, 67.0] | 0.0 |
| BMI | (median [IQR]) | 29.2 | [25.5, 34.6] | 0.0 |
| Population | (n (%)) |  |  | 0.0 |
| African |  | 832 | (13.4) |  |
| Asian |  | 223 | (3.6) |  |
| European |  | 4010 | (64.5) |  |
| Hispanic |  | 519 | (8.3) |  |
| Other |  | 636 | (10.2) |  |
| Sex (male) | (mean (SD)) | 0.381 | (0.486) | 0.0 |
| T01 IPK | (mean (SD)) | 0.051 | (0.068) | 0.0 |
| T02 UMU | (mean (SD)) | 0.052 | (0.074) | 0.0 |
| T03 IMG | (mean (SD)) | 0.047 | (0.061) | 0.0 |
| T04 IVO | (mean (SD)) | 0.054 | (0.068) | 0.0 |
| T05 HVK | (mean (SD)) | 0.050 | (0.090) | 0.0 |
| T06 BMV | (mean (SD)) | 0.040 | (0.061) | 0.0 |
| T07 GGG | (mean (SD)) | 0.050 | (0.069) | 0.0 |
| T08 DDD | (mean (SD)) | 0.064 | (0.111) | 0.0 |
| T09 PPM | (mean (SD)) | 0.064 | (0.093) | 0.0 |
| T10 BMK | (mean (SD)) | 0.037 | (0.055) | 0.0 |
| T11 IIL | (mean (SD)) | 0.036 | (0.072) | 0.0 |
| T12 IIC | (mean (SD)) | 0.059 | (0.073) | 0.0 |
| T13 PFV | (mean (SD)) | 0.075 | (0.099) | 0.0 |
| T14 BGM | (mean (SD)) | 0.054 | (0.084) | 0.0 |
| T15 OOO | (mean (SD)) | 0.060 | (0.090) | 0.0 |
| T16 PBM | (mean (SD)) | 0.054 | (0.074) | 0.0 |
| T17 VKL | (mean (SD)) | 0.049 | (0.083) | 0.0 |
| T18 ILL | (mean (SD)) | 0.044 | (0.067) | 0.0 |
| T19 SSS | (mean (SD)) | 0.060 | (0.092) | 0.0 |
| AHI | (median [IQR]) | 17.5 | [10.2, 29.0] | 0.0 |
| T90 | (median [IQR]) | 0.1 | [0.0, 1.6] | 0.1 |
| ArBdn | (median [IQR]) | 126.4 | [94.0, 168.6] | 0.0 |
| ArThr | (median [IQR]) | 123.1 | [112.3, 136.7] | 0.5 |
| ChxDly | (median [IQR]) | 12.5 | [10.8, 14.7] | 0.3 |
| HypBdn | (median [IQR]) | 18.1 | [8.4, 37.1] | 0.1 |
| HRBdn | (median [IQR]) | 132.3 | [72.2, 236.6] | 0.3 |
| LG1 | (median [IQR]) | 0.5 | [0.4, 0.6] | 0.3 |
| LGN | (median [IQR]) | 0.4 | [0.3, 0.5] | 0.3 |
| VntBdn | (median [IQR]) | 269.7 | [140.4, 499.6] | 0.0 |
| Vpassive | (median [IQR]) | 96.0 | [93.3, 97.8] | 0.0 |
| Vactive | (median [IQR]) | 101.5<sup>1</sup> | [97.8, 103.8] | 0.1 |
| Vcomp | (median [IQR]) | 4.6 | [3.2, 6.7] | 0.1 |

<sup>1</sup> Vactive, Vcomp, and Vpassive are reported on the natural eupnea-normalized scale and can exceed 100% in patients with greater-than-eupneic ventilation during sleep; rank-normalized values were used in regression analyses (Methods).

### ATM Topics

Age-dependent topic modeling (ATM) was employed in 36,426 ascertained OSA cases (80% training samples and 20% validation samples, Methods). Model input consisted of Phecode diagnoses along with age of first diagnosis, derived from EHR after quality control procedures via validated algorithmic ascertainment methods (Methods). We selected 305 phecodes for ATM training: of 354 codes meeting prevalence thresholds (Methods), 305 showed Bonferroni-significant association with OSA case status (p < 0.05 / 354) in logistic regression adjusting for sex, age, BMI, self-reported ancestry, and healthcare utilization. EHR data was limited to first-available diagnoses among EHR encounters at age 25 or older, due to inconsistent availability of childhood EHR.

After model training, the best-performing model was carried forward to association testing in the held-out independent testing dataset (Methods). The selected final model identified 19 distinct topics (see Table 2a and Supplementary Figures), capturing a diverse range of OSA comorbidities, including cardiometabolic, respiratory, neurological, and psychiatric diagnoses. For the selected model, age-dependent multimorbidity trajectories across the life-course are defined by disease-topic weights that vary in a continuous non-linear fashion with age, fitted using splines with three knots (Supplementary Figures).

**Table 2a.**
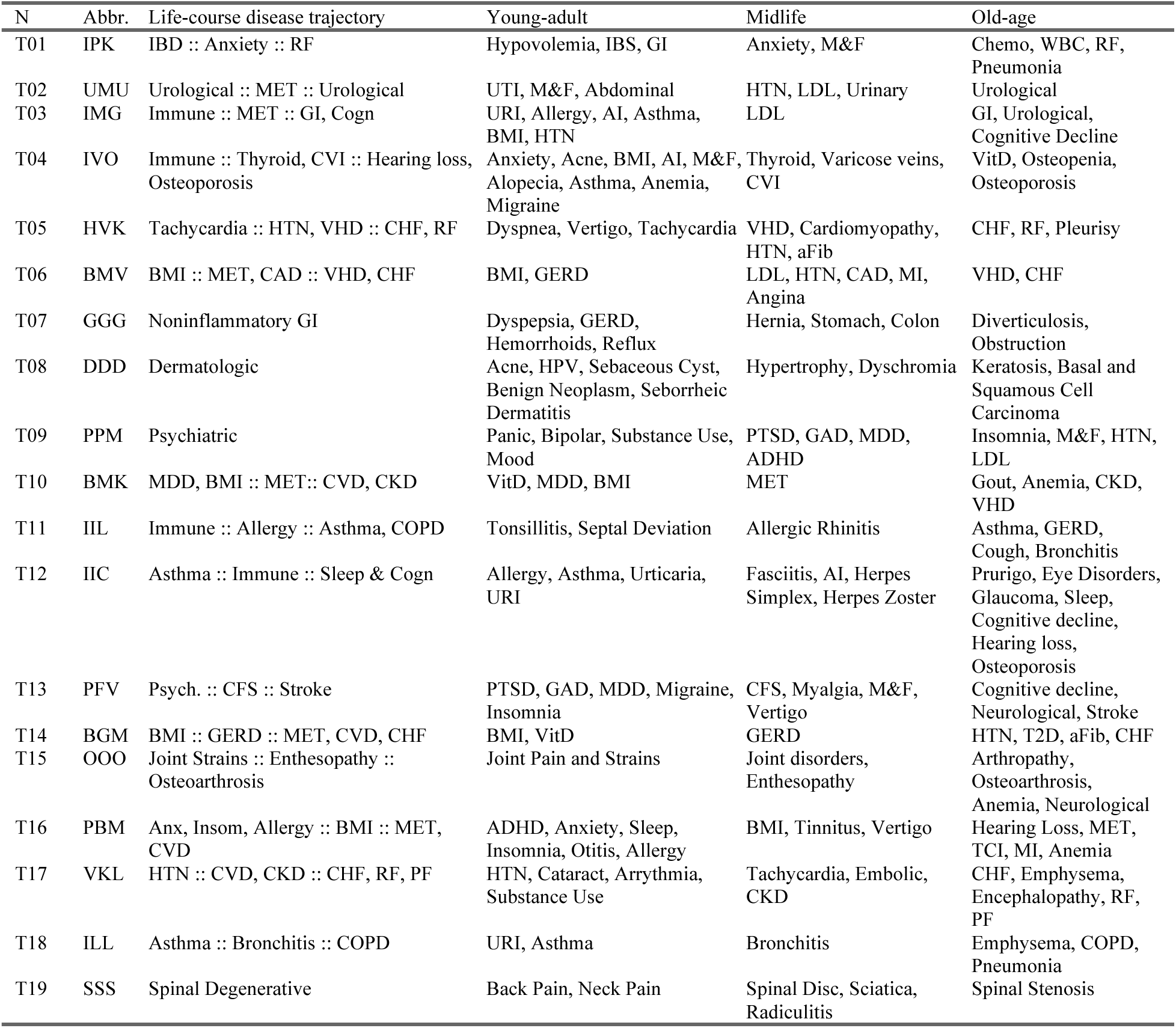
Description of MGB ATM topics. ATM topic shorthand abbreviations (Abbr.): B: high BMI/obesity, C: dementia/cognitive, D: dermatologic, F: chronic fatigue-related, G: gastrointestinal, H: cardiac arrhythmias and myelopathies, I: immune-mediated, K: renal, L: pulmonary, M: metabolic, O: orthopedic/osteological, P: psychiatric, S: spinal-degenerative, U: urological, V: vascular/cardiovascular/cerebrovascular. Life-course disease trajectory: Young-adult (∼25-45 years of age) :: Mid-life (45–65) :: Old-age (65+). Abbreviations for disorders: ADHD: attention deficit hyperactivity disorder; aFib: atrial fibrillation; AI: autoimmune; Anx: anxiety; BMI: body mass index; CAD: coronary artery disease; CFS: chronic fatigue syndrome; Chemo: chemotherapy; CHF: congestive heart failure; CKD: chronic kidney disease; Cogn: cognitive decline; CVD: cardiovascular disease; CVI: chronic venous insufficiency; GAD: generalized anxiety disorder; GERD: gastroesophageal reflux disease; GI: gastrointestinal; HPV: human papillomavirus; HTN: hypertension; IBS: irritable bowel syndrome; Immune: immune-mediated; Insom: insomnia; LDL: low-density lipoprotein cholesterol; MDD: major depressive disorder; MET: metabolic syndrome; M&F: malaise and fatigue; MI: myocardial infarction; Mood: mood disorder; Neuro: neurological disorders; PF: pulmonary failure; PTSD: post-traumatic stress disorder; Reflux: acid reflux; RF: acute renal failure; Sleep: sleep disorders; Spinal Disc: spinal disc disorder; Substance Use: substance use disorder; TCI: transient cerebral ischemia; T2D: type 2 diabetes; URI: upper respiratory infection; UTI: urinary tract infection; VHD: valvular heart disease; VitD: vitamin D deficiency; WBC: elevated white blood cell count.

| N | Abbr. | Life-course disease trajectory | Young-adult | Midlife | Old-age |
| --- | --- | --- | --- | --- | --- |
| T01 | IPK | IBD :: Anxiety :: RF | Hypovolemia, IBS, GI | Anxiety, M&F | Chemo, WBC, RF, Pneumonia |
| T02 | UMU | Urological :: MET :: Urological | UTI, M&F, Abdominal | HTN, LDL, Urinary | Urological |
| T03 | IMG | Immune :: MET :: GI, Cogn | URI, Allergy, AI, Asthma, BMI, HTN | LDL | GI, Urological, Cognitive Decline |
| T04 | IVO | Immune :: Thyroid, CVI :: Hearing loss, Osteoporosis | Anxiety, Acne, BMI, AI, M&F, Alopecia, Asthma, Anemia, Migraine | Thyroid, Varicose veins, CVI | VitD, Osteopenia, Osteoporosis |
| T05 | HVK | Tachycardia :: HTN, VHD :: CHF, RF | Dyspnea, Vertigo, Tachycardia | VHD, Cardiomyopathy, HTN, aFib | CHF, RF, Pleurisy |
| T06 | BMV | BMI :: MET, CAD :: VHD, CHF | BMI, GERD | LDL, HTN, CAD, MI, Angina | VHD, CHF |
| T07 | GGG | Noninflammatory GI | Dyspepsia, GERD, Hemorrhoids, Reflux | Hernia, Stomach, Colon | Diverticulosis, Obstruction |
| T08 | DDD | Dermatologic | Acne, HPV, Sebaceous Cyst, Benign Neoplasm, Seborrheic Dermatitis | Hypertrophy, Dyschromia | Keratoses, Basal and Squamous Cell Carcinoma |
| T09 | PPM | Psychiatric | Panic, Bipolar, Substance Use, Mood | PTSD, GAD, MDD, ADHD | Insomnia, M&F, HTN, LDL |
| T10 | BMK | MDD, BMI :: MET :: CVD, CKD | VitD, MDD, BMI | MET | Gout, Anemia, CKD, VHD |
| T11 | IIL | Immune :: Allergy :: Asthma, COPD | Tonsillitis, Septal Deviation | Allergic Rhinitis | Asthma, GERD, Cough, Bronchitis |
| T12 | IIC | Asthma :: Immune :: Sleep & Cogn | Allergy, Asthma, Urticaria, URI | Fasciitis, AI, Herpes Simplex, Herpes Zoster | Prurigo, Eye Disorders, Glaucoma, Sleep, Cognitive decline, Hearing loss, Osteoporosis |
| T13 | PFV | Psych. :: CFS :: Stroke | PTSD, GAD, MDD, Migraine, Insomnia | CFS, Myalgia, M&F, Vertigo | Cognitive decline, Neurological, Stroke |
| T14 | BGM | BMI :: GERD :: MET, CVD, CHF | BMI, VitD | GERD | HTN, T2D, aFib, CHF |
| T15 | OOO | Joint Strains :: Enthesopathy :: Osteoarthritis | Joint Pain and Strains | Joint disorders, Enthesopathy | Arthropathy, Osteoarthritis, Anemia, Neurological |
| T16 | PBM | Anx, Insom, Allergy :: BMI :: MET, CVD | ADHD, Anxiety, Sleep, Insomnia, Otitis, Allergy | BMI, Tinnitus, Vertigo | Hearing Loss, MET, TCI, MI, Anemia |
| T17 | VKL | HTN :: CVD, CKD :: CHF, RF, PF | HTN, Cataract, Arrhythmia, Substance Use | Tachycardia, Embolic, CKD | CHF, Emphysema, Encephalopathy, RF, PF |
| T18 | ILL | Asthma :: Bronchitis :: COPD | URI, Asthma | Bronchitis | Emphysema, COPD, Pneumonia |
| T19 | SSS | Spinal Degenerative | Back Pain, Neck Pain | Spinal Disc, Sciatica, Radiculitis | Spinal Stenosis |

**Table 2b.** Selected ATM Model Performance. Model performance for the final selected ATM model, showing prediction odds ratios for this model in the training, validation, and test data sets. OR_top1: Odds ratio comparing the odds that a patient’s true next diagnosis ranks within the top 1% of ATM-predicted diseases versus the odds it ranks within the top 1% most prevalent diseases. OR_top10: Odds ratio comparing the odds that a patient’s true next diagnosis ranks within the top 10% of ATM-predicted diseases versus the odds it ranks within the top 10% most prevalent diseases. Individual age-specific disease risk is computed from individualized ATM topic scores by linear combination with the topic’s age-specific disease loadings (Methods). Values >1 indicate ATM outperforms naive prevalence-based prediction.

| Cohort | # Subjects | OR_top1 | OR_top10 |
| --- | --- | --- | --- |
| Training (OSA cases) | 29,042 | 1.43 | 1.57 |
| Validation (OSA cases) | 7,276 | 1.43 | 1.55 |
| Test (OSA Case-Control) | 34,384 | 1.50 | 1.61 |
| Test (OSA cases) | 11,689 | 1.42 | 1.55 |
| Test (OSA controls) | 22,695 | 1.58 | 1.66 |
| Test (PSG) | 6,220 | 1.56 | 1.52 |
| Test (PSG, AHI≥15) | 3,594 | 1.45 | 1.52 |
| Test (PSG, AHI<15) | 2,626 | 1.79 | 1.52 |

Individualized ‘topic risk-scores’ were calculated from longitudinal age-of-incidence data derived from the EHR, providing a summary of individuals’ life-course comorbidity patterns, as well as predicting future incident comorbidities. ATM optimizes the prediction of incident diagnoses by grouping diagnoses into clusters, using co-occurring diagnoses associated with specific ages, and linking these to later incident diagnoses, resulting in a set of ATM topics representing distinct life-course multimorbidity trajectories. The 19 ATM topic risk-scores were successful in predicting incident diagnoses across training, validation, and testing cohorts, having similar prediction OR (see Methods for description of this prediction performance metric) across subgroups and OSA case status (Table 2b).

### Associations Between ATM Topics and OSA Case-Control Status

We tested associations between each of the 19 age-dependent comorbidity progression topics and OSA case-control status using separate linear regression models (Table 3) in the OSA case-control testing dataset (11,689 OSA cases, 22,695 matched controls). All 19 topics showed statistically significant associations with OSA status. Each model treated an ATM topic as the dependent variable and OSA status as the exposure, adjusting for age, sex, BMI, self-reported ancestry, and healthcare utilization (number of EHR encounters and age at first EHR encounter). To simplify interpretation and protect against type-I error, we standardized ATM topics via rank normalization (RN), such that effect sizes (β) represent standardized mean differences in topic scores between OSA cases and controls, adjusting for covariates. Below, we highlight the four topics with the largest effect sizes (β ≥ 0.3), indicating substantial differences between OSA cases and controls; see Table 3 for complete results.

**Table 3:** ATM Topic associations with OSA status. Association with OSA status for each of the 19 ATM topics. N: sample size. R^2^: model R-square. Beta: OSA association effect size, Pr(>|t|): OSA association p-value, Bonf: association passes Bonferroni threshold on 19 tests, FDR-adj p: False-discovery-rate adjusted p-value, FDR: passes FDR threshold of 0.05. Topic-Disease/Disorder Abbreviations (Abbr.): B: high BMI/obesity, C: cognitive decline and dementia, D: dermatologic, F: chronic fatigue, G: gastrointestinal, H: cardiac arrhythmias and myelopathies, I: immune-mediated, K: kidney, L: lung, M: metabolic, O: orthopedic/connective/osteological, P: psychiatric, R: rheumatic, S: spinal-degenerative, U: urological, V: cardiovascular/cerebrovascular. Life-course disease trajectory: Early-adulthood (∼25-45 years of age) :: Mid-life (45–65) :: Late-adulthood (65+).

| Topic # | Topic Abbr. | Life-course disease trajectory | Exposure | N | R<sup>2</sup> | Beta | P-value |
| --- | --- | --- | --- | --- | --- | --- | --- |
| T01 | IPK | IBD :: Anxiety :: RF | OSA | 34384 | 0.046 | -0.089 | 5.7e- 17 |
| T02 | UMU | Urological :: MET :: Urological | OSA | 34384 | 0.036 | 0.15 | 3.1e- 45 |
| T03 | IMG | Immune :: MET :: GI, Cogn | OSA | 34384 | 0.051 | 0.31 | 1.1e-175 |
| T04 | IVO | Immune :: Thyroid, CVI :: Hearing, Osteoporosis | OSA | 34384 | 0.13 | 0.17 | 3.1e- 61 |
| T05 | HVK | Tachycardia :: HTN, VHD :: CHF, RF | OSA | 34384 | 0.053 | 0.20 | 2.9e- 75 |
| T06 | BMV | BMI :: MET, CAD :: VHD, CHF | OSA | 34384 | 0.075 | 0.15 | 8.4e- 45 |
| T07 | GGG | Noninflammatory GI | OSA | 34384 | 0.043 | 0.16 | 1.1e- 48 |
| T08 | DDD | Dermatologic | OSA | 34384 | 0.12 | 0.30 | 3.8e-180 |
| T09 | PPM | Psychiatric | OSA | 34384 | 0.080 | 0.33 | 1.3e-206 |
| T10 | BMK | MDD, BMI :: MET:: CVD, CKD | OSA | 34384 | 0.078 | 0.19 | 1.9e- 69 |
| T11 | IIL | Immune :: Allergy :: Asthma, COPD | OSA | 34384 | 0.12 | 0.28 | 1.9e-156 |
| T12 | IIC | Asthma :: Immune :: Sleep & Cogn | OSA | 34384 | 0.10 | 0.16 | 2.9e- 52 |
| T13 | PFV | Psych. :: CFS :: Stroke | OSA | 34384 | 0.063 | 0.19 | 2.0e- 70 |
| T14 | BGM | BMI :: GERD :: MET, CVD, CHF | OSA | 34384 | 0.13 | 0.25 | 4.0e-134 |
| T15 | OOO | Joint Strains :: Enthesopathy :: Osteoarthritis | OSA | 34384 | 0.056 | 0.058 | 1.3e- 7 |
| T16 | PBM | Anx, Insom, Allergy :: BMI :: MET, CVD | OSA | 34384 | 0.040 | 0.30 | 2.6e-171 |
| T17 | VKL | HTN :: CVD, CKD :: CHF, RF, PF | OSA | 34384 | 0.065 | -0.061 | 1.4e- 8 |
| T18 | ILL | Asthma :: Bronchitis :: COPD | OSA | 34384 | 0.033 | 0.086 | 9.6e- 16 |
| T19 | SSS | Spinal Degenerative | OSA | 34384 | 0.027 | 0.062 | 1.1e- 8 |

Specifically, the following age-dependent comorbidity progression patterns associated strongly with OSA status:

**Psychiatric (PPM):** progressing from panic disorder, bipolar disorder, substance use, and mood disorders (early adulthood), to post-traumatic stress disorder (PTSD), generalized anxiety disorder (GAD), major depressive disorder (MDD), and attention-deficit/hyperactivity disorder (ADHD) (middle age), to insomnia, mixed neurocognitive features, hypertension, and elevated LDL cholesterol (older age) (β = 0.33; p = 1.3e-206)
**Immune-Metabolic to Gastrointestinal and Cognitive Disorders (IMG):** progressing from upper respiratory infections, allergies, autoimmune disease, asthma, elevated BMI, and hypertension (early adulthood), to hyperlipidemia (middle age), to gastrointestinal and urological disorders, and cognitive decline (older age) (β = 0.31; p = 1.1e-175)
**Dermatologic (DDD)**: progressing from acne, human papillomavirus (HPV), sebaceous cysts, benign skin neoplasms, and seborrheic dermatitis (early adulthood), to skin hypertrophy and dyschromia (middle age), to actinic keratosis, basal cell carcinoma, and squamous cell carcinoma (older age) (β = 0.30; p = 3.8e-180)
**Psychiatric-Metabolic with Sensory and Vascular Sequelae (PBM):** progressing from attention-deficit/hyperactivity disorder (ADHD), anxiety, adjustment reactions, sleep disturbance, insomnia, otitis media, and allergic conditions (early adulthood), to obesity, tinnitus, and vertigo (middle age), to hearing loss, metabolic dysfunction, transient cerebral ischemia (TCI), myocardial infarction (MI), and anemia (older age) (β = 0.30; p = 2.6e-171)

### Associations Between ATM Topics and OSA-related PSG measures

Descriptions of the PSG exposures are provided for reference (Table 4a). Association analysis of ATM topics with continuous PSG-derived OSA endophenotypes revealed significant relationships between multiple topics and specific PSG measures (Table 4). The analysis adjusted for the same covariates used in OSA case-control testing and again employed rank normalization (RN) standardization of topic scores. PSG measures were log transformed or rank normalized, as stated in Table 4a. Significance was declared for 0.05 false discovery rate (FDR-adjusted p-value < 0.05), across 247 tests (19 ATM topics and 13 PSG measures).

**Table 4a.** ATM Topic associations per PSG endophenotype. Summary of model-based associations relating ATM topics (dependent variable) with PSG measures (independent variable), showing each the 13 PSG measures, along with their statistical transformation, description, number of associations and ATM topics associated (ordered by p-value, increasing). See Supplementary Table 2 for full PSG association results. Topic-Disease/Disorder Abbreviations (Abbr.): B: high BMI/obesity, C: cognitive decline and dementia, D: dermatologic, F: chronic fatigue, G: gastrointestinal, H: cardiac arrhythmias and myelopathies, I: immune-mediated, K: kidney, L: lung, M: metabolic, O: orthopedic/connective/osteological, P: psychiatric, R: rheumatic, S: spinal-degenerative, U: urological, V: cardiovascular/cerebrovascular. FDR: false-discovery rate. Trans.: Transformation: data transformation when used as exposure in association regression model: logarithmic (LOG), rank normalization (RN) *ATM topics that were associated with the given PSG measure at FDR 0.05.

| # | PSG Measure | Trans. | Description | # Assoc* | ATM Topics Associated* |
| --- | --- | --- | --- | --- | --- |
| 01 | AHI | LOG | Apnea Hypopnea Index (AHI3p) | 4 | PFV(-), BMK(+), IMG(+), PPM(-) |
| 02 | T90 | RN | Total Sleep Time Spent at SpO2 < 90% | 10 | VKL(+), ILL(+), PBM(-), DDD(-), HVK(+), PFV(-), IIC(-), IVO(-), UMU(-), BMV(+) |
| 03 | ArBdn | LOG | Arousal Burden | 2 | IVO(-), PFV(-) |
| 04 | ArThr | LOG | Arousal Threshold | 3 | BMK(+), IMG(+), PFV(-) |
| 05 | ChxDly | LOG | Chemoreflex Delay | 4 | VKL(-), BMK(-), ILL(-), SSS(+) |
| 06 | HypBdn | LOG | Hypoxic Burden | 5 | PFV(-), BMK(+), IMG(+), PPM(-), PBM(-) |
| 07 | HRBdn | LOG | Heart Rate Burden | 7 | PFV(-), IMG(+), PPM(-), VKL(-), HVK(-), BMK(+), IPK(-) |
| 08 | VntBdn | LOG | Ventilatory Burden | 6 | PFV(-), BMK(+), IMG(+), PPM(-), IPK(-), PBM(-) |
| 09 | LG1 | RN | Loop Gain (1 cycle/min) | 3 | BMK(+), VKL(+), IIL(-) |
| 10 | LGN | RN | Loop Gain (natural frequency) | 3 | ILL(-), OOO(-), HVK(+) |
| 11 | Vpassive | RN | Patency: Passive Conditions | 7 | PFV(+), IMG(-), BMK(-), ILL(+), IPK(+), PPM(+), VKL(+) |
| 12 | Vactive | RN | Patency: Active Conditions | 7 | VKL(+), ILL(+), HVK(+), PPM(+), IPK(+), PFV(+), BMV(+) |
| 13 | Vcomp | RN | Pharyngeal Dilator Activation | 5 | VKL(+), PPM(+), HVK(+), ILL(+), BMV(+) |

**Table 4b.**
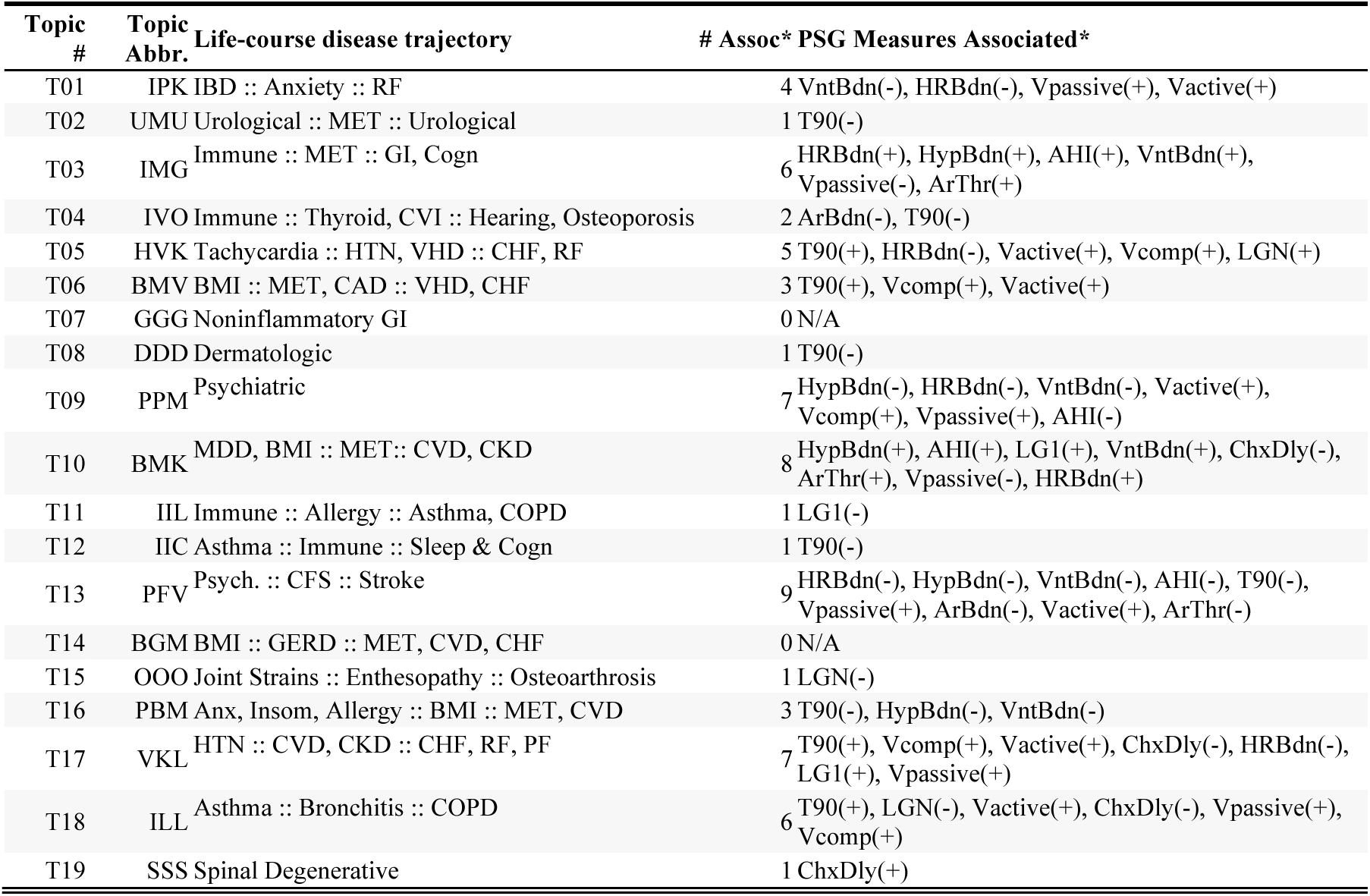
PSG endophenotype associations per ATM Topic. Summary of model-based associations relating ATM topics (dependent variable) with PSG measures (independent variable), showing each the 19 ATM topics, along with their description, number of associations and associated PSG measures (ordered by p-value, increasing). See Supplementary Table 2 for full PSG association results. *PSG measures that were associated with the given ATM topic at FDR 0.05. N/A: not available (no associations at FDR 0.05). FDR: false-discovery rate.

**Table 4c.**
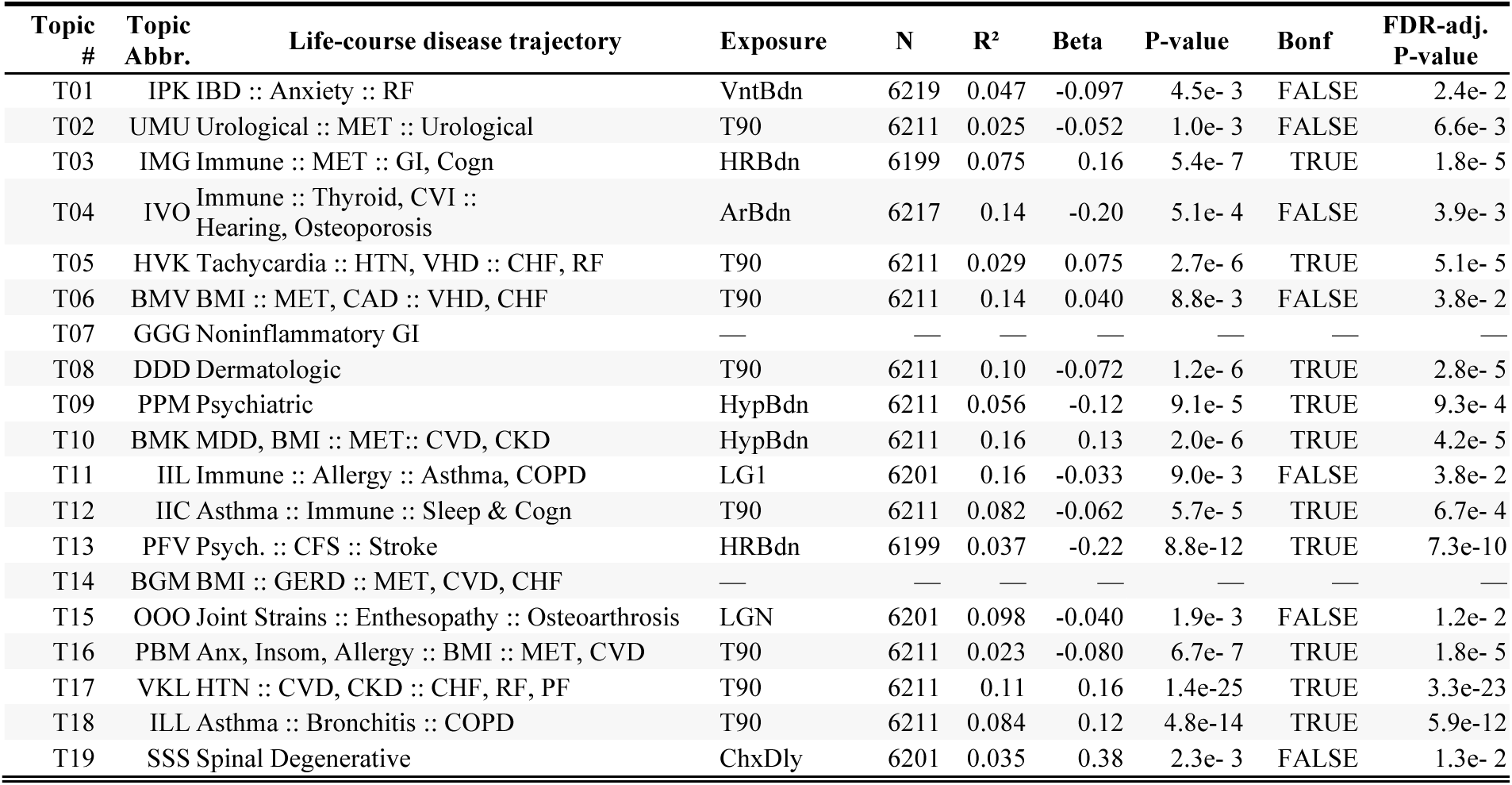
Selected endophenotypes associated with ATM Topics. Selected associations between ATM Topics and PSG measures: we report the PSG measure with the strongest evidence of association (smallest p-value), among those associations at FDR 0.05. Complete statistically significant (Bonferroni and FDR) results are given in Supplementary Table 2. FDR: false-discovery rate. N/A: not available (no associations at FDR 0.05).

For brevity, in Table 4c, among results with less than 0.05 false discovery rate (FDR-adjusted p-value < 0.05), we report the PSG measures with the strongest statistical evidence per ATM topic; complete FDR-significant results presented in Supplementary Table 2. Considering positive associations, hypoxic burden was the exposure with strongest statistical evidence of association with the Topic BMK (Early adulthood high BMI, progressing to midlife metabolic disorders, and late life kidney disease, i.e., BMI/obesity, Metabolic, Kidney). Likewise, T90 was the exposure most strongly associated with Topics HVK (Heart, Vascular, Kidney), ILL (Immune-mediated, Lung, Lung), and VKL (Vascular, Kidney, Lung), heart rate burden with IMG (Immune-mediated, Metabolic, Gastrointestinal), and chemoreflex delay with SSS (Spondylosis/Degenerative Spinal).

Several topics’ strongest associations with PSG measures were via inverse associations, indicating more severe hypoxemia, autonomic dysregulation, or other PSG measures associated with lower risk of incident disease for a given topic. These negative associations included Topics DDD (Dermatologic), IIC (Immune-mediated, Immune-mediated, Cognitive), PBM (Psychiatric, BMI/obesity, Metabolic), and UMU’s (Urological, Metabolic, Urological) inverse relationship with T90, Topic IVO’s (Immune-mediated, Vascular, Orthopedic) inverse relationship with arousal burden, Topic PFV’s (Psychiatric, Fatigue, Vascular) inverse relationship with heart rate burden, and Topic PPM’s (Psychiatric, Psychiatric, Metabolic) inverse relationship with hypoxic burden, Topic IPK’s (Immune-mediated, Psychiatric, Kidney) inverse relationship with ventilatory burden, Topic IIL (Immune-mediated, Immune-mediated, Lung) and OOO’s (Orthopedic) respectively with LG1 and LGN (loop gain at 1 cycle/min and at the natural frequency, respectively).

At FDR < 0.05 there were many additional positive associations (Supplementary Table 2) as well as inverse associations between Topics BMK and VKL with chemoreflex delay, Topic HVK and heart rate burden, and Topic ILL with LGN.

### Sensitivity Analyses

We performed two sensitivity analyses to evaluate robustness of our findings. First, we restricted analyses to patients meeting the clinical definition of OSA (AHI ≥15; n=3,588 to n=3,594). Second, in the full PSG testing dataset, we implemented LASSO regression with post-selection inference (Methods), allowing simultaneous adjustment for all 13 PSG measures. Both analyses yielded results qualitatively consistent with our primary findings. In the OSA-restricted analysis, 28 of 66 FDR-significant associations (42.4%) from the primary analysis remained at FDR<0.05 (see Supplementary Table 3). Among primary FDR-significant associations, effect estimates showed mainly modest changes (median: −11.5%; IQR: −33.0% to +13.5%) with high correlation in effect size between analyses (Spearman ρ=0.96). One association (Topic PPM with Vpassive) showed directional reversal but was no longer FDR-significant. The LASSO analysis (Supplementary Table 4) identified 14 of 66 primary FDR-significant associations that retained FDR<0.05 significance after mutual adjustment. Effect estimates showed substantial shrinkage (median: −57.6%; IQR: −89.1% to −12.0%) with moderate correlation to primary results (Spearman ρ=0.65). The regularization process set 12 associations to zero; an additional 40 were estimated but no longer FDR-significant. Five associations showed directional reversals, of which one (Topic IMG with ventilatory burden) retained FDR significance (β = +0.13 in primary versus β = −0.08 in LASSO). With this exception, no associations retained FDR significance with opposite sign in either sensitivity analysis, supporting overall directional consistency with primary findings.

## DISCUSSION

Age-dependent topic modeling of longitudinal EHR data from OSA patients identified 19 distinct comorbidity trajectories, all significantly associated with OSA status in independent testing data, of which 17 also associated with at least one PSG-derived physiological measure. These findings demonstrate that life-course comorbidity patterns in OSA are structured, predictive, and linked to measurable differences in OSA pathophysiology.

These results provide a framework to connect a patient’s observed comorbidity history with both physiological characteristics and future disease outcomes. OSA patients do not follow a single comorbidity trajectory; rather, distinct patterns of age-dependent disease incidence associate with different PSG profiles and different late-life endpoints. To illustrate we offer three clinical scenarios that show how integrating comorbidity trajectory information with PSG findings could inform individualized OSA management.

### Metabolic trajectories and OSA severity

Topics IMG and BMK represent metabolic comorbidity trajectories that associate with more severe measures of OSA physiology. Both topics showed positive associations with AHI, hypoxic burden, arousal threshold, and ventilatory burden (Table 4). IMG, progressing from early-adulthood immune and inflammatory conditions through midlife metabolic dysfunction to late-adulthood gastrointestinal disease and cognitive decline, also associated with heart rate burden, a measure of apnea-associated autonomic response. BMK, progressing from early-adulthood depression and obesity through midlife metabolic syndrome to late-adulthood cardiovascular and chronic kidney disease, showed the same associations as IMG including heart rate burden, and additionally associated with chemoreflex delay (inverse) and loop gain. Elevated loop gain reflects ventilatory instability common in individuals with cardiac and cerebrovascular disease. These findings extend prior work linking OSA to inflammatory dysregulation (*1*), cardiometabolic and kidney disease (*32, 33*), as well as neurocognitive decline and dementia (*34–36*), by characterizing age-related incidence patterns in these trajectories. Additionally, the findings reveal specific OSA endophenotypes, indicative of autonomic dysregulation and intermittent hypoxia, having the strongest association with each trajectory. Notably, restricting to the AHI ≥ 15 subset, the effect sizes for BMK’s associations with hypoxic burden, AHI, and ventilatory burden approximately doubled (Supplementary Table 3), consistent with a dose-response relationship and with prior reports linking hypoxic burden to incident cardiovascular and metabolic disease (*37*). These findings are consistent with a model in which autonomic dysregulation and intermittent hypoxia mediate the relationship between OSA and downstream cardiometabolic, renal, and neurocognitive outcomes (*1, 2, 10*). To establish this more definitively, statistical mediation analysis would ideally involve longitudinal endophenotype measurements, which were not available in this study. Clinically, observing early metabolic risk factors and inflammatory conditions in an OSA patient along with the above endophenotypes may signify future risk for metabolic syndrome, and potential benefit from monitoring for specific late-life endpoints predicted by each trajectory: cognitive and gastrointestinal disease for IMG, cardiovascular and renal disease for BMK. It will require further research to determine if specific endophenotype profiles, for example high heart rate burden, are prognostic for IMG over BMK outcomes.

### Inverse associations between psychiatric trajectories and burdens

Psychiatric comorbidity topics PPM and PBM showed among the highest increased scores in cases vs controls, indicating that age-specific diagnoses in psychiatric comorbidity trajectories are more common in OSA populations. Yet both topics showed inverse associations with hypoxic burden and ventilatory burden, and PPM additionally showed inverse associations with AHI and heart rate burden (Table 4), while PPM also showed positive associations with all three airway patency measures (Vactive, Vcomp, Vpassive), suggesting a less collapsible airway. The combination of positive association with OSA status and inverse association with specific physiological burdens suggests that psychiatric comorbidity is prominent in OSA yet that patients with higher psychiatric comorbidity have less severe physiological burdens and lower airway collapsibility. Of note, the psychiatric topic PFV showed inverse associations with both arousal threshold and arousal burden among the psychiatric trajectories (Table 4b). The lower arousal threshold indicates greater arousability, while at the same time a lower arousal burden, may reflect shorter respiratory-event-driven arousals, or fewer arousals. Further investigation of arousal architecture in this trajectory is warranted.

In the AHI ≥ 15 sensitivity analysis, the inverse associations with hypoxic burden and heart rate burden remained directionally consistent but were modestly attenuated and no longer FDR significant. This pattern contrasts with the cardiometabolic trajectories, where the same sensitivity analysis showed strengthened PSG metric associations. Restricting to AHI ≥ 15 removes patients most likely to have been referred for insomnia rather than for OSA, so the consistent direction but partial attenuation of the inverse psychiatric–burden associations is consistent with collider bias contributing to but not fully explaining this result. In contrast, the airway patency associations were largely attenuated and no longer statistically significant in the AHI ≥ 15 subgroup.

After accounting for referral bias, remaining inverse associations between psychiatric trajectories and physiologic burdens could be explained by anxiety or insomnia in OSA associating with lower BMI and consequently lower hypoxic and ventilatory burden. The psychiatric topics are expected to encompass a subgroup with unrecognized comorbid insomnia and sleep apnea (COMISA), which is increasing acknowledged as a unique clinical entity (*38*). These overall findings would be consistent with a primary role for systemic inflammation, associated with both OSA and with anxiety and insomnia, linking OSA to psychiatric comorbidity independent of hypoxic burden and high BMI. Patients in these topic groups may show low risk based on a severity assessment that relies primarily on burden measures previously associated with a cardiometabolic risk profile, but may still be at risk for psychiatric outcomes and later-life cardiovascular outcomes. These findings suggest that psychiatric-trajectory OSA patients, including the subgroup with (COMISA), may benefit from integrated psychiatric and sleep management and potential monitoring for inflammatory markers alongside traditional OSA severity measures. Additionally, PPM and PBM diverge on late-life endpoints, with PPM predictive of neurocognitive features and hypertension and PBM indicating risk of future cardiovascular events, which may necessitate careful monitoring and follow-up for indicators of the specific psychiatric comorbidity profile.

### Early-onset versus late-onset asthma as distinct OSA respiratory phenotypes

Topics IIC and ILL both involve asthma and are positively associated with OSA case status, but differ in age of onset and show divergent PSG profiles. IIC, representing early-onset asthma with immune-mediated conditions progressing to cognitive decline, showed an inverse association with T90 (higher hypoxemia associated with lower IIC comorbidity). ILL, representing late-onset asthma progressing to COPD, showed the opposite: a strong positive association with T90 (along with positive associations with airway patency measures and inverse associations with loop gain and chemoreflex delay). This directional contrast maps onto the established clinical distinction between allergic early-onset asthma, driven by inflammatory and genetic mechanisms, and non-allergic late-onset asthma, linked to metabolic dysfunction and impaired pulmonary mechanics (*39–42*). The positive association of T90 with ILL is consistent with impaired lung function contributing to nocturnal hypoxemia, while the inverse association in IIC suggests that early-onset asthma-OSA link may operate differently, e.g. through shared inflammatory pathways. The absence of COPD in IIC in contrast with ILL may account for the apparent ‘protection’ against severe hypoxemia indicated by its negative association with T90. Clinically, this implies late-onset asthma in an OSA patient should prompt attention to pulmonary function and nocturnal hypoxemia, while early-onset asthma should prompt monitoring for inflammatory markers as well as later cognitive decline.

### Cautious interpretation

The associations reported here require careful interpretation with respect to causality. Topic trajectories represent predictive patterns linking co-occurring diagnoses, not necessarily causal chains; for example, GERD in Topic BGM may share metabolic risk factors with later heart disease rather than causing it. Similarly, positive relationships between T90 and topics involving heart failure and renal failure (BMV, HVK, VKL) likely reflect reverse or bidirectional causation, where impaired cardiac or pulmonary function manifests as increased nocturnal hypoxemia (*43*). Additionally, ATM topic scores represent the probability that a subject’s incident diagnoses will come from that topic, meaning that they are compositional and must sum to one across topics for each individual; as such, elevated scores on some topics necessarily reduce scores on others. The two inverse case-control associations (IPK and VKL) should be interpreted in this context, as OSA cases preferentially load onto specific comorbidity trajectories with consequent relative depletion in others. In light of these considerations our results should be considered hypothesis-generating rather than indicating definitive causal relationships.

### Additional findings and interpretation

Several ATM topics representing well-established OSA comorbidity patterns provided confirmatory evidence for our approach, including metabolic-cardiovascular trajectories (BGM, BMK) and inflammatory-respiratory trajectories (IIC, IIL, ILL), detecting known OSA associations while further characterizing nuanced age-specific patterns in sequential incident diagnoses. Beyond these expected patterns, ATM also identified associations with comorbidity domains not typically surveyed in OSA cohort studies, notably dermatologic disease (Topic DDD), potentially linked through systemic inflammation, oxidative stress, and intermittent hypoxia (*44*), as well as degenerative spinal conditions (Topic SSS), potentially linked through systemic inflammation and obesity-related mechanical stress (*45*). A further advantage of the ATM framework is to highlight comorbidity patterns that appear similar in early-adulthood but diverge in later trajectories: early inflammatory disorders branch into respiratory (IIL), cognitive (IIC), or metabolic-gastrointestinal (IMG) endpoints, while early psychiatric symptoms diverge into metabolic-vascular (PBM) or mood-cognitive (PPM) trajectories. These divergence points suggest specific monitoring periods or intervention windows may exist where targeted therapies could redirect patients toward more favorable outcomes. Across several of these trajectories, including IMG and IIC, we hypothesize that intermittent hypoxemia exacerbates systemic inflammation (*46*), with later divergence potentially reflecting interactions between this shared risk pathway and individual genetic predispositions or life-course exposures.

### Strengths and limitations

We analyzed a large real-world sample using both clinical polysomnography and incident diagnoses inferred from longitudinal EHR, linked with clinical measurements and demographic covariates. We employed recommended best practices for EHR analysis, including using validated phenotyping algorithms (MAP, PheVis) to process raw EHR data and more accurately determine incident diagnoses, and adjusted models for healthcare utilization. The ATM approach offers key advantages for studying OSA multimorbidity: it captures age-dependent temporal structure in comorbidity patterns, it generates heritable and generalizable phenotypes as previously demonstrated in UK Biobank and All of Us (*25*), and it condenses analysis of hundreds of diagnostic codes into clinically interpretable topics while reducing multiple testing burden.

This work has several limitations. First, in an open EHR system, we cannot fully confirm when diagnostic codes represent disease incidence. Second, EHR completeness may vary between OSA cases and controls and could introduce bias; to mitigate this we adjusted for total healthcare encounters and age at first encounter, however, unadjusted differences may remain. Third, ATM captures the evolution of comorbidity trajectories over the life course as a single score per individual, but it still may be of interest whether ATM profiles change in response to OSA incidence, treatment, or disease progression. However, we were unable to do so reliably in the current data due to expected diagnostic delay (*47*) and limited availability of longitudinal OSA data. Fourth, prediction performance of future age-specific comorbidity incidence was slightly higher in controls than cases, possibly reflecting greater variability in comorbidity age of incidence among cases related to varying OSA age-of-onset or progression; however, prediction performance was largely similar across cohorts. Finally, as a data-driven modeling approach, ATM may yield different topic sets in different datasets, though these have been previously shown to be correlated across cohorts (*25*).

### Summary

Our results demonstrate the utility of ATM for characterizing OSA-associated multimorbidity patterns. By identifying predictive life-course comorbidity trajectories linked to both OSA status and PSG-derived physiological measures, this approach provides a framework for characterizing OSA heterogeneity from real-world data. The associations revealed are interpretable and suggest potential applications for clinical risk stratification in OSA patients based on age-specific comorbidity patterns and PSG endophenotype characteristics, as illustrated by the metabolic-renal, psychiatric, and respiratory scenarios discussed above. Future research should attempt validation in additional populations, as well as incorporate longitudinal measures of OSA severity and treatment response to determine whether effective OSA treatment can modify these comorbidity trajectories and whether risk stratification via ATM can improve clinical outcomes.

## MATERIALS AND METHODS

### Study Design

This study aimed to characterize patterns of comorbidity progression associated with obstructive sleep apnea (OSA) and their physiological correlates, using a retrospective observational design based on electronic health records (EHR) and clinical polysomnography (PSG) data from the Mass General Brigham (MGB) health system. The primary objectives were to (1) identify distinct longitudinal comorbidity trajectories using age-dependent topic modeling (ATM), (2) evaluate whether these patterns differ between OSA cases and matched controls, and (3) examine associations between inferred comorbidity trajectories and PSG-derived physiological traits (endophenotypes). The analysis leveraged existing phenotyping pipelines previously developed within the MGB system. This included standardized EHR preprocessing, mapping ICD9 and ICD10 codes to the standardized ‘phecode’ clinical ontology, along with phenotype classification using Multimodal Automated Phenotyping (MAP), and OSA case ascertainment using the PheVis algorithm, followed by 2:1 propensity score matching to define a case-control cohort. ATM was trained on the subset of identified OSA cases, excluding those with either PSG data or genetic data. ATM topics were then projected into two independent testing datasets: a matched case-control sample and a PSG cohort. The PSG cohort included patients with diagnostic or split-night PSG recordings, with endophenotypes derived from preprocessed signals data.

### Patient Selection and Data Sources

We analyzed electronic health records from adults (≥18 years) in the Mass General Brigham health system using the Research Patient Data Registry, a clinical data warehouse spanning 10 hospitals in greater Boston, MA, USA (*48*). The study received IRB approval (protocols 2024P002360, 2021P003546, 2010P001765). EHR data were extracted on March 6, 2023 for the main cohort and January 9, 2025 for patients with genetic or polysomnographic data, so these dates represent end of EHR follow-up.

Clinical data included ICD-9 and ICD-10 diagnosis codes mapped to Phecodes (*49*) and free-text clinical notes. Potential OSA cases had at least one phecode for sleep apnea (327.3) or obstructive sleep apnea (327.32); controls had neither. Polysomnographic data came from Faulkner Hospital sleep clinic, including patients regardless of OSA status; when multiple studies existed, we used the first available chronologically. Genotyping availability was extracted from the MGB Biobank (*50, 27*). We excluded patients for incomplete covariates (e.g. BMI) or for failing to meet a minimal amount of healthcare utilization (“data floor”; ≥3 diagnoses, ≥3 clinical encounters, and ≥3 notes)(*51*).

### EHR Preprocessing and OSA Phenotyping

OSA cases were ascertained using the PheVis algorithm (*30*), which combines diagnostic codes with clinical note terms to improve identification beyond Phecode-based selection. Note terms were extracted using cTAKES, coded to the UMLS ontology, and filtered to OSA-relevant terms from select OSA literature via the SAFE procedure as detailed elsewhere (*52*). The algorithm was validated against chart reviews of 484 patients evaluated by sleep clinicians using ICSD-3 criteria (*11*). Patients with AHI ≥15 from sleep study reports or clinical note extraction (excluding machine-based or stage-specific values) were additionally classified as true cases. Age at diagnosis was the date of first OSA phecode or sleep study; BMI used measurements closest to this date.

Controls were matched 2:1 to cases using propensity score matching with hard thresholds for BMI (±5 kg/m²), and birth year (±5 years). Matching variables included birth year (to adjust for temporal changes in EHR coding), BMI, self-reported race/ethnicity (mapped to continental ancestry categories: African, Asian, European, Hispanic, Other), and binned healthcare encounters (*53*), with separate matching by sex. Control age was set to the matched case’s diagnosis age (*51*). Some cases had incomplete matching when fewer than two eligible controls were available.

### Training and testing cohorts

To ensure valid statistical testing and preserve nominal Type-1 error, the training and testing cohorts for ATM were pre-defined to be strictly non-overlapping. The ATM testing dataset consisted of two partly overlapping sub-groups: A) OSA cases with genotyping or genetic sequence data along with their matched controls, and B) patients with available clinical PSG from Faulkner Hospital sleep clinic (including OSA cases, controls or ambiguous). The ATM training dataset consisted of the remaining identified OSA cases, after excluding all those in the testing subset.

### ATM Topics

Age-dependent Topic Modeling (ATM) was employed to identify age-dependent clusters or trajectories patterning comorbidity cooccurrence observed in the longitudinal EHR data. The model uses ‘topics’ to represent static ‘trait-like’ correlated patterns in comorbidity occurrence across the lifespan and allows individuals’ next observed EHR code (Phecode) to be predicted from their estimated topic weightings, which are assumed to be fixed over time. Phecodes analyzed by ATM were limited to those associated with OSA, after applying prevalence filters and removing nonspecific phecodes. Phecodes ascertained via the MAP algorithm were required to have ≥1% prevalence in both males and females in both the RPDR case-control cohort and the Faulkner PSG cohort. Among these phecodes only those with ≥2% prevalence among RPDR OSA cases (combined males and females) were used to train the ATM model. Ambiguous non-specific codes were removed (Phecodes 1005: “Other symptoms”; 1010: “Other tests”; 1019: “Other ill-defined and unknown causes of morbidity and mortality”; 1100: “Family history”; and related 1XXX codes). Among the 354 Phecodes passing these filters, we tested for association with OSA case status using logistic regression in the non-held-out RPDR cohort (training and validation subsets combined, including cases and matched controls), adjusting for birth year, BMI, sex, self-reported ancestry, and healthcare utilization. Phecodes with Bonferroni-significant association (p < 0.05/354) were selected for ATM modeling.

The ATM model was then trained and topic risk scores were calculated in the testing dataset, via the following three-step process.

1. **Model Fitting**: Per algorithm authors’ recommendation, ATM models were fit to the EHR in 80% of the training dataset. Models were fit in the ATM open-source *R* package, across a range of parameters. Specifically, the number of topics was varied between 10 and 20, and the flexibility of the age-dependent pattern of EHR diagnosis was varied (linear, polynomial, or spline with 1, 2 or 3 knots). For each set of model parameters, variational inference methods were used to estimate topic weights for each individual and the age-dependent topic loadings for each Phecode.
2. **Model Validation and Selection**: Among the ATM models fit across varying parameters, model selection was performed in the EHR validation sample consisting of the remaining 20% of the training dataset, based on optimizing model prediction performance (according to the default recommended prediction odds ratio statistic).
3. **Calculating ATM topic ‘risk scores’:** The final trained ATM model was applied to the independent testing datasets to generate individualized topic scores. For each patient, EHR diagnosis codes and their corresponding ages of first diagnosis were input into the pre-trained model, to compute posterior topic scores reflecting the degree of alignment with each of the 19 age-dependent comorbidity trajectories. This calculation was performed separately for the matched case-control testing cohort and the PSG cohort from Faulkner Hospital. The resulting topic scores represent fixed, trait-like summaries of each patient’s comorbidity profile and were used as dependent variables in subsequent association analyses with OSA case status and PSG-derived endophenotypes.

### PSG Variables

PSG metrics related to OSA severity and pathophysiology were computed from signals data according to previously described algorithms. (*11, 52, 54*):

| # | Abbr. | Metric (Units) | Definition |
| --- | --- | --- | --- |
| 1 | AHI | Apnea-hypopnea index (events/h) | Apnea-hypopnea index, recomputed from PSG signals data. Apneas: >90% airflow reduction from pre-event baseline for $\geq 10$ s; hypopneas: $\geq 30\%$ airflow reduction with $\geq 3\%$ desaturation or arousal. Rate of events per hour of sleep. (54) |
| 2 | T90 | Sleep Time with $\text{SpO}_2 < 90\%$ (% of sleep time) | Percentage of total sleep time with oxyhemoglobin saturation below 90%, recomputed from oximetry signals data. (54) |
| 3 | ArBdn | Arousal burden (% of sleep time) | Cumulative duration of all cortical arousals, expressed as a fraction of total sleep time. Captures combined arousal frequency and duration per arousal. (55) |
| 4 | ArThr | Arousal threshold (% eupnea, square-root transformed, centered at 100%) | Mean ventilatory drive immediately prior to scored EEG arousals, estimated per 7-minute window from the chemoreflex feedback control model. Lower values reflect greater arousability from sleep. (54, 56) |
| 5 | ChxDly | Chemoreflex delay (s) | Delay parameter of the fitted chemoreflex feedback control model, representing the latency between reduced ventilation and the rise in ventilatory drive. (54, 57) |
| 6 | HypBdn | Hypoxic burden<br>(%/min/h) | Respiratory event-specific area under the oxyhemoglobin desaturation curve measured from pre-event baseline, summed over events and normalized to total sleep time. Captures cumulative respiratory event-related hypoxemia, per hour of sleep, as combined frequency, SpO2 depth, and duration. (58) |
| 7 | HRBdn | Heart rate burden,<br>(bpm/h) | Respiratory event-specific pulse rate response, measured from pre-event baseline, total respiratory event-related change in heart rate (delta heart rate), summed over events, and normalized to total sleep time. Captures cumulative elevated heart rate from respiratory events, per hour of sleep. (37) |
| 8 | VntBdn | Ventilatory burden<br>(% of breaths) | Respiratory event-specific area under the mean-normalized ventilation signal, summed over scored events and normalized to total sleep time. Captures cumulative ventilatory deficit from respiratory events, per hour of sleep. (59) |
| 9 | LG1 | Loop gain at 1<br>cycle/min<br>(dimensionless) | Sensitivity of the ventilatory feedback loop to a 1 cycle/min disturbance: the magnitude of the ventilatory drive response to a specific ventilatory perturbation. Reflects the product of chemoreflex sensitivity and plant gain. Summarized as the median across 7-min windows. (54, 57) |
| 10 | LGN | Overall loop gain<br>(dimensionless) | Overall ventilatory instability representing predisposition to cyclic central sleep apnea. Combined effect of chemoreflex sensitivity and chemoreflex delay, evaluated at the natural system frequency. Summarized as the median across 7-min windows. (54, 57) |
| 11 | Vpassive | Entilation at passive<br>(eupneic) drive<br>(% eupnea) | Median ventilation at normal (eupneic) ventilatory drive, derived from the decile-binned ventilation-drive relationship with linear interpolation between bins. Lower Vpassive reflects greater anatomical collapsibility of the pharyngeal airway. (54, 60) |
| 12 | Vactive | Ventilation at active<br>ventilatory drive<br>(% eupnea) | Median ventilation at the ventilatory drive corresponding to the arousal threshold, i.e., maximal drive during sleep, estimated from the decile-binned ventilation-drive relationship. (54, 60) |
| 13 | Vcomp | Pharyngeal dilator<br>activation,<br>compensation (%<br>eupnea) | Vactive minus Vpassive: the increase in ventilation between eupneic drive and maximal drive, reflecting pharyngeal dilator muscle responsiveness. (54, 60) |

### Statistical Analysis of ATM Topic Associations

We conducted association analyses between ATM-derived comorbidity topic scores and two classes of exposures: 1) OSA case-control status and 2) PSG-derived endophenotypes. For both analyses, each topic score was treated as a continuous dependent variable in a separate linear regression model. To facilitate interpretation and ensure standardized effect sizes across topics, ATM topic scores were transformed using rank normalization (RN) prior to analysis. As described above, caliper matching was conducted to best utilize the available sample, while ensuring common support in covariate distributions across OSA cases and controls, avoiding extrapolation beyond the data in modeling. However, to further protect against confounding all models were adjusted for the matching covariates. Adjustment covariates consisted of age at PSG or OSA diagnosis, sex, BMI, and self-reported ancestry, as well as measures of healthcare utilization (age at first EHR encounter and total number of encounters).

In case-control analyses, the primary exposure was OSA status (case vs. matched control). In PSG analyses, each of the 13 PSG-metrics was modeled separately as the exposure of interest. To correct for multiple comparisons across the 19 ATM topics and 13 PSG metrics (247 tests), a Bonferroni-adjusted significance threshold of p < 2.0 × 10⁻⁴ was used. All associations meeting a false discovery rate (FDR) < 0.05 were considered for reporting in the manuscript. PSG measures were rank-normalized or log-transformed prior to analysis, depending on their distributional characteristics, to meet model assumptions and mitigate influence of outliers (transformations listed in Table 4a). All analyses were conducted in R (version 4.4.2) using the renv package for reproducible dependency management. ATM topic modeling used the ATM package (version 0.0.0.9000; github.com/Xilin-Jiang/ATM, SHA: 1163a77). Key statistical packages included glmnet (version 4.1-8) for LASSO regularization, selectiveInference (version 1.2.5) for post-selection inference in sensitivity analyses, and survival (version 3.8-3) for time-to-event analyses.

Analyses used complete case samples, excluding subjects with missing values for any covariate. For case-control and OSA cohort analyses, required covariates included age, BMI, sex, self-reported ancestry, age at first EHR diagnosis, and total healthcare encounters. For PSG cohort analyses, AHI was additionally required. Covariate missingness was minimal (<5% for most variables) and assumed to be missing completely at random.

### Sensitivity Analyses

To assess the potential impact of Berkson’s bias in the PSG testing cohort, we performed a subset sensitivity analysis. In clinical practice, referral for PSG often arises from divergent pathways—either OSA-specific symptoms and risk factors (e.g., snoring, obesity, hypertension), or complaints of insomnia or hypersomnolence potentially linked to psychiatric conditions. This dual referral pattern may result in collider bias, particularly in analyses involving psychiatric comorbidity topics, by inducing spurious inverse associations between OSA severity and psychiatric disease. To mitigate this, we repeated all PSG topic association analyses within the AHI ≥15 subgroup to examine whether topic-endophenotype associations were robust to exclusion of individuals with lower AHI from PSG data.

As a second sensitivity analysis, we performed multi-exposure LASSO regression to evaluate whether ATM topic associations persisted after mutual adjustment for correlated PSG variables. For each ATM topic, we fit a LASSO model including all 13 PSG metrics simultaneously, along with the same covariates used in primary analyses. The regularization parameter (λ) was selected via 10-fold cross-validation using λ_min (minimizing prediction error). Because standard inference is invalid after variable selection, we applied post-selection inference using the fixedLassoInf() function from the selectiveInference R package to obtain valid p-values conditional on the selected model. Importantly, LASSO regularization applies to all predictors including covariates, not just exposures; multi-exposure results therefore represent which variables show independent associations when all are considered together, rather than PSG effects controlling for fixed covariates.

To systematically assess robustness of findings across sensitivity analyses, we compared effect estimates from primary single-exposure models with those from sensitivity analyses using multiple concordance metrics. Spearman’s rank correlation (ρ) was calculated for all comparisons to assess monotonic association between primary and sensitivity estimates, with Pearson correlation (r) as a secondary metric. For each association, we calculated percent change in effect size ((Sensitivity estimate – Primary estimate) / Primary estimate × 100%). Among FDR-significant primary findings, we classified sensitivity results into mutually exclusive categories indicating directional consistency. Sign changes were also tabulated separately for FDR-significant results.

## Data Availability

All data for present study are available upon reasonable request to the authors, given appropriate institutional approvals.

## Acknowledgments

The authors wish to thank the Mass General Brigham Research Patient Data Registry (RPDR) for providing the data in an analyzable format, as well as the patients who participated and the staff of the Brigham and Women’s Faulkner Hospital sleep lab.

## Author contributions

Conceptualization: BEC

Methodology: BEC, MOG, RA

Investigation: BEC, MOG, RA

Visualization: MOG

Funding acquisition: BEC

Project administration: BEC

Supervision: BEC

Writing – original draft: MOG

Writing – review & editing: All coauthors

## Data and materials availability

The primary EHR, demographic, and polysomnography data contain protected health information and cannot be shared publicly. Access to these data will be considered upon request to the corresponding author, potentially requiring execution of a data use agreement and material transfer agreement (MTA), subject to Mass General Brigham IRB approval and institutional review. Summary-level results for the primary and sensitivity analyses are provided in the supplementary materials. Analysis code is available at https://github.com/MatthewOGoodman/ATM_OSA_Analysis. The ATM software package is publicly available at https://github.com/Xilin-Jiang/ATM.

## Supplement

### Supplementary Figures

**Fig S1.**
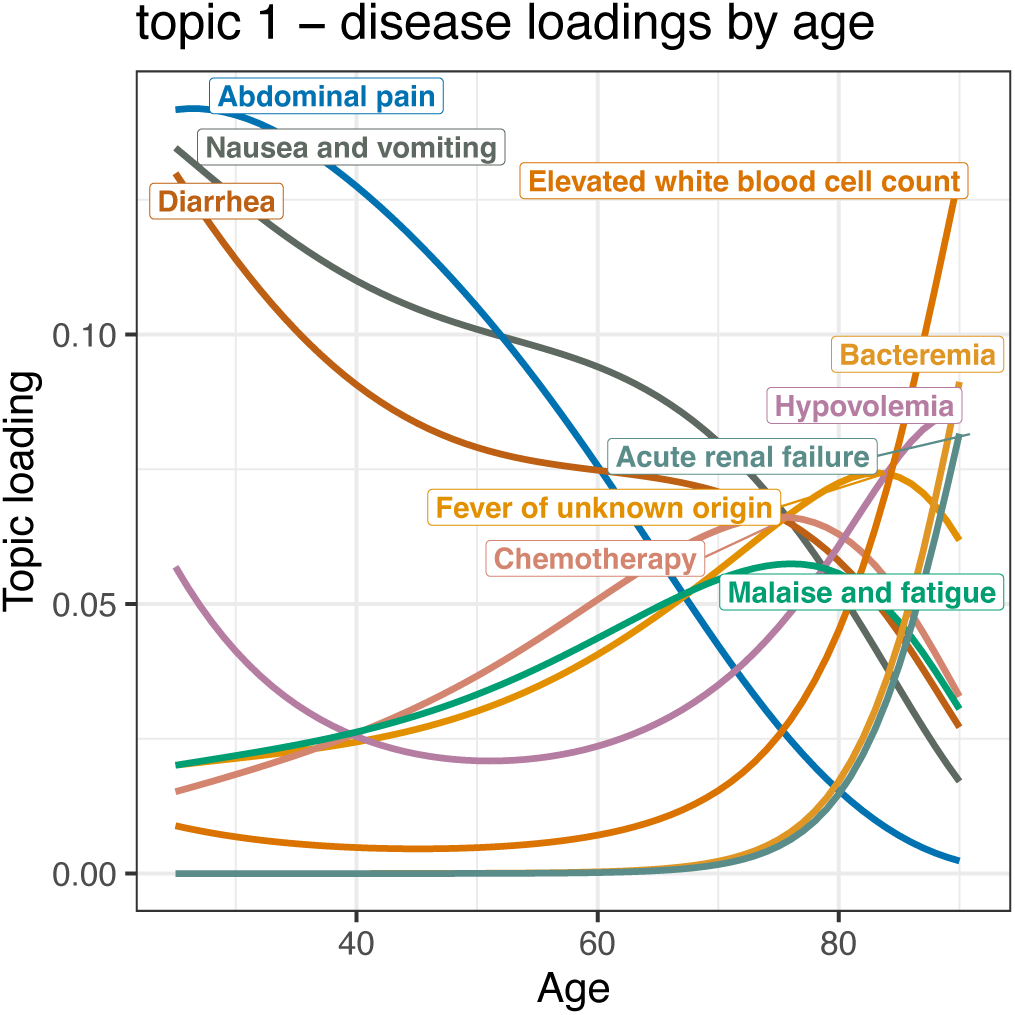
ATM Disease-Topic loadings: T01 IPK.

**Fig S2.**
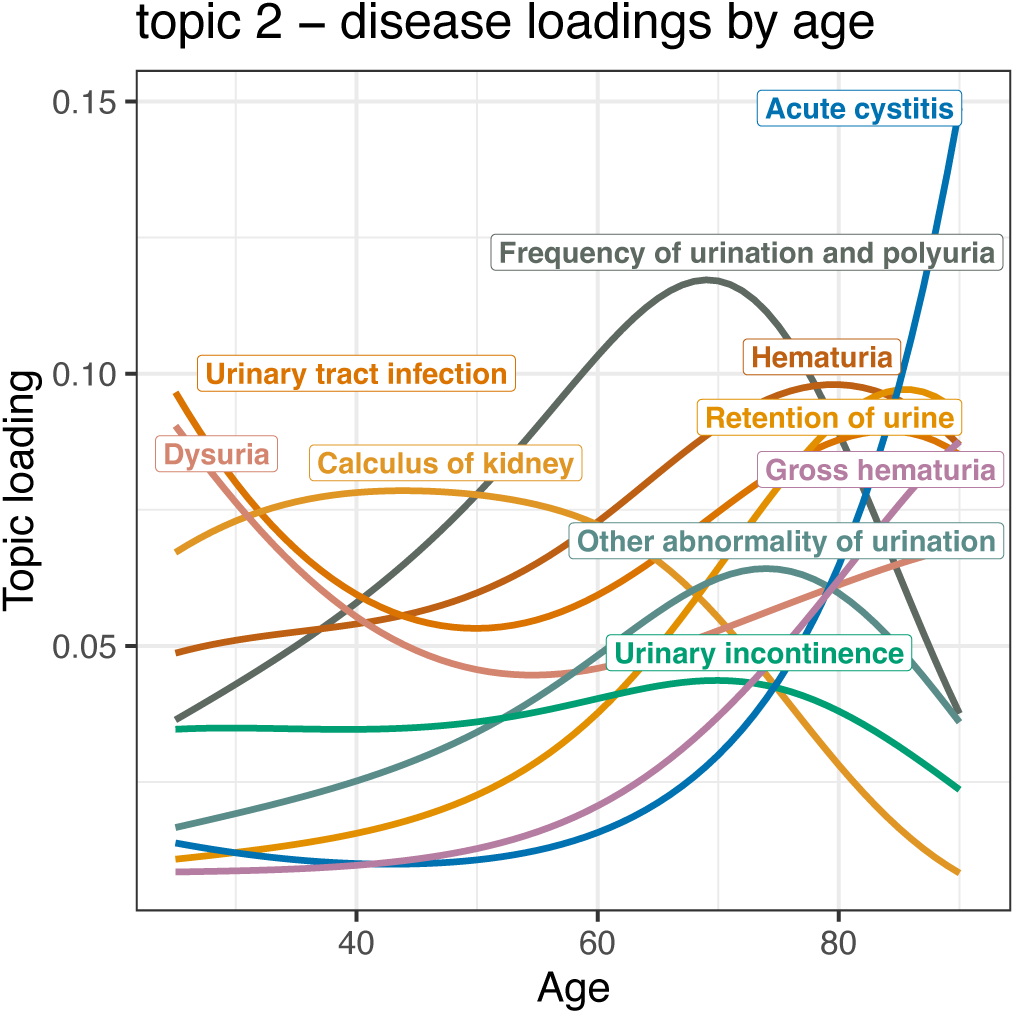
ATM Disease-Topic loadings: T02 UMU.

**Fig S3.**
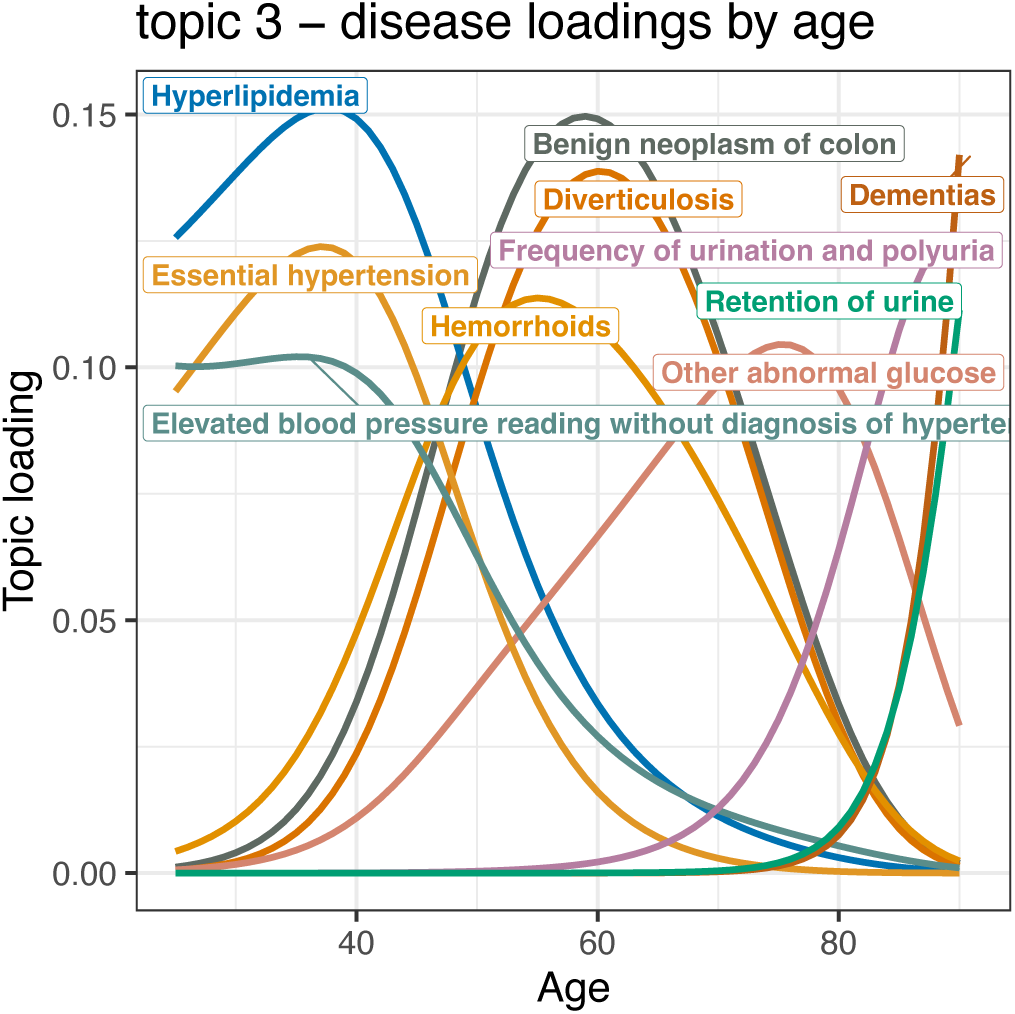
ATM Disease-Topic loadings: T03 IMG.

**Fig S4.**
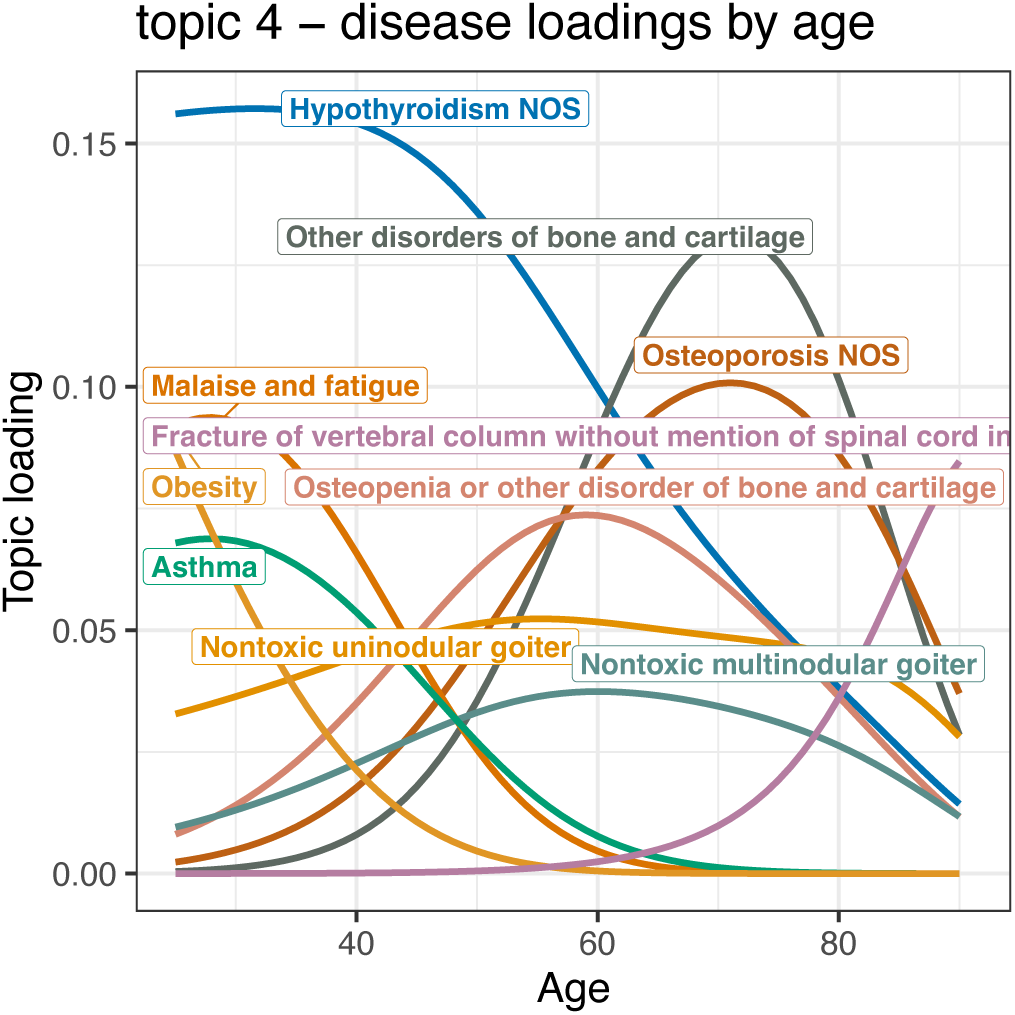
ATM Disease-Topic loadings: T04 IVO.

**Fig S5.**
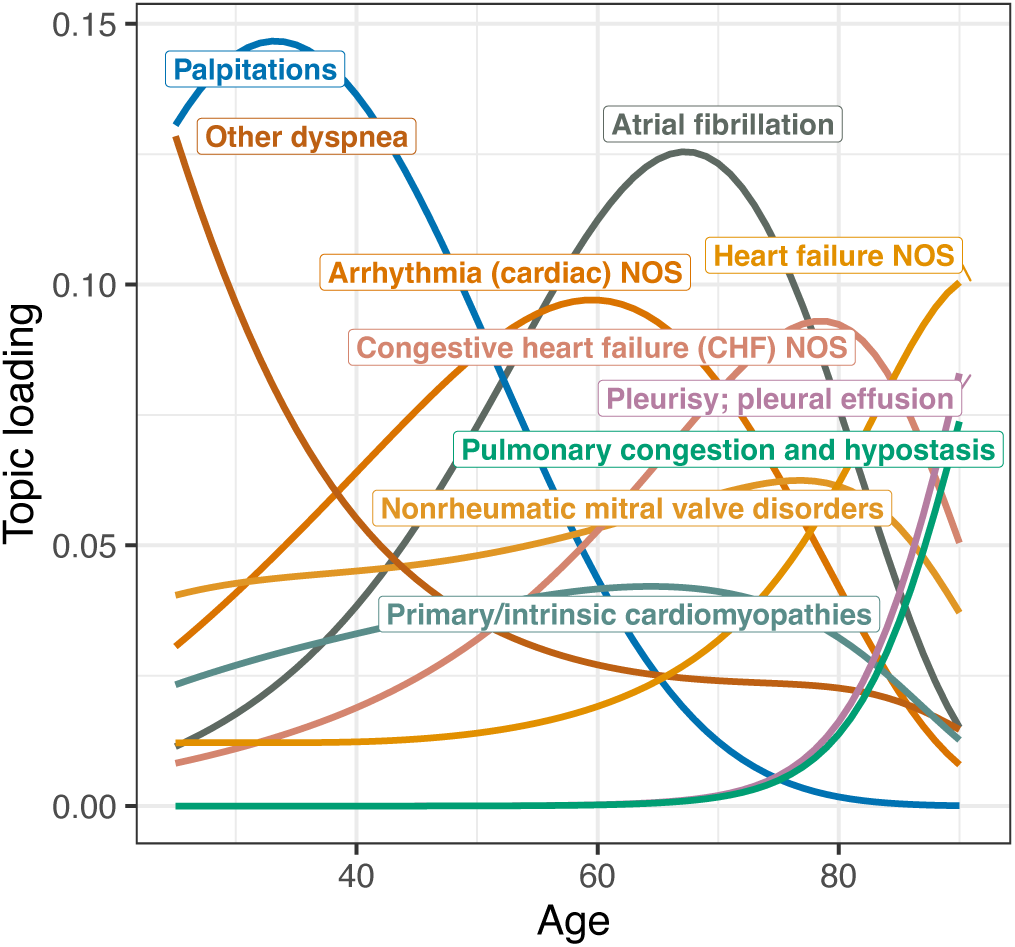
ATM Disease-Topic loadings: T05 HVK.

**Fig S6.**
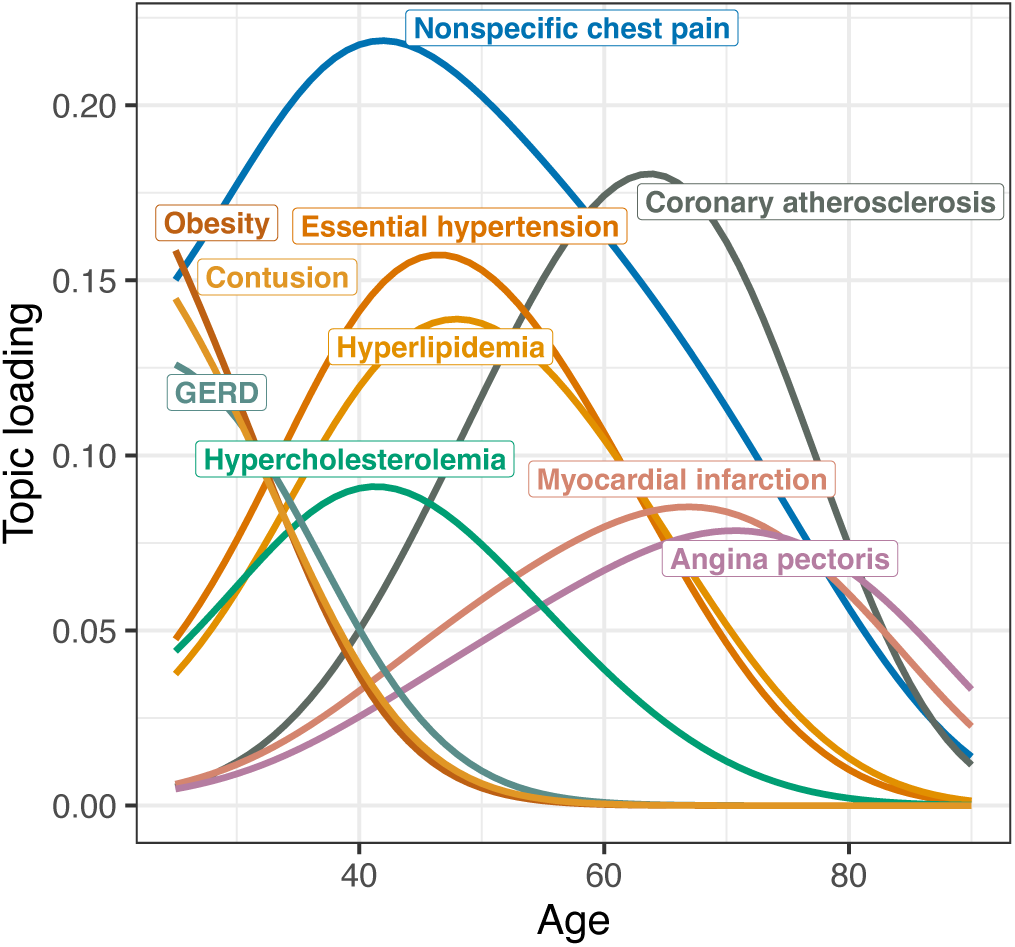
ATM Disease-Topic loadings: T06 BMV.

**Fig S7.**
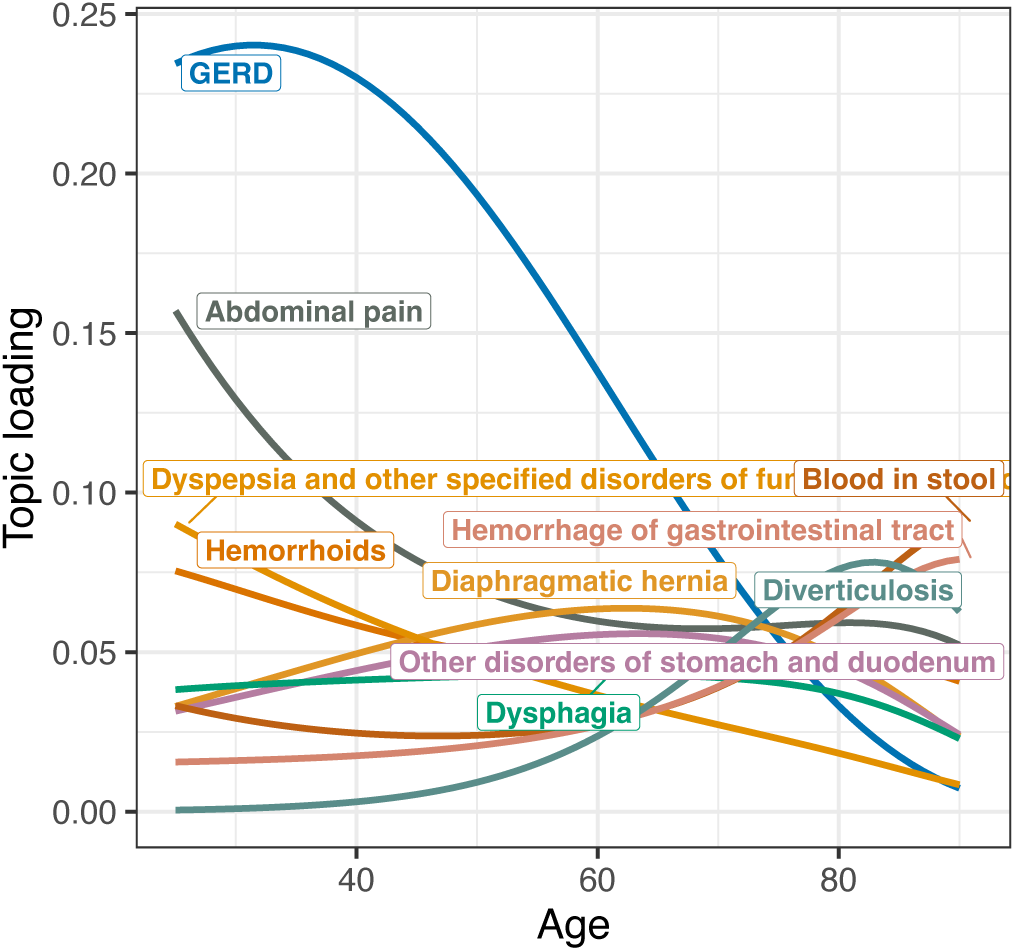
ATM Disease-Topic loadings: T07 GGG.

**Fig S8.**
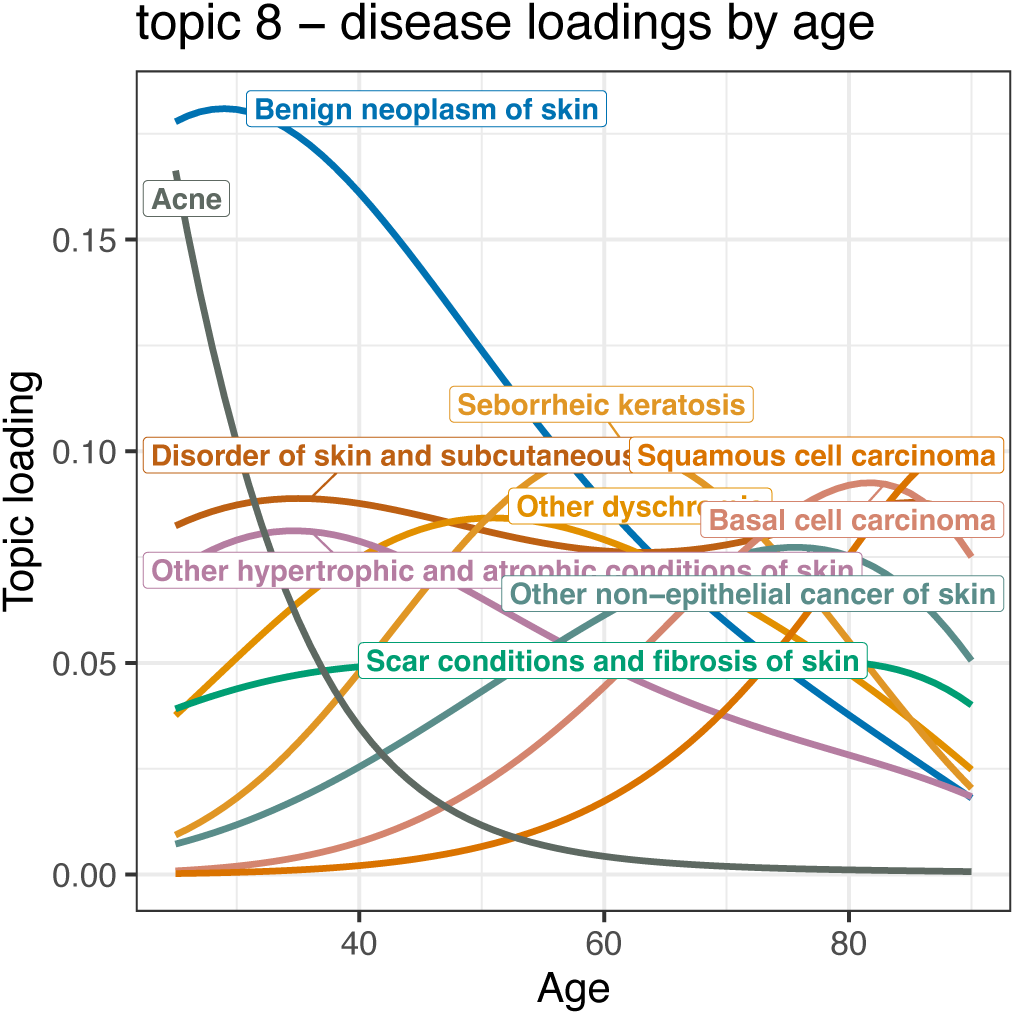
ATM Disease-Topic loadings: T08 DDD.

**Fig S9.**
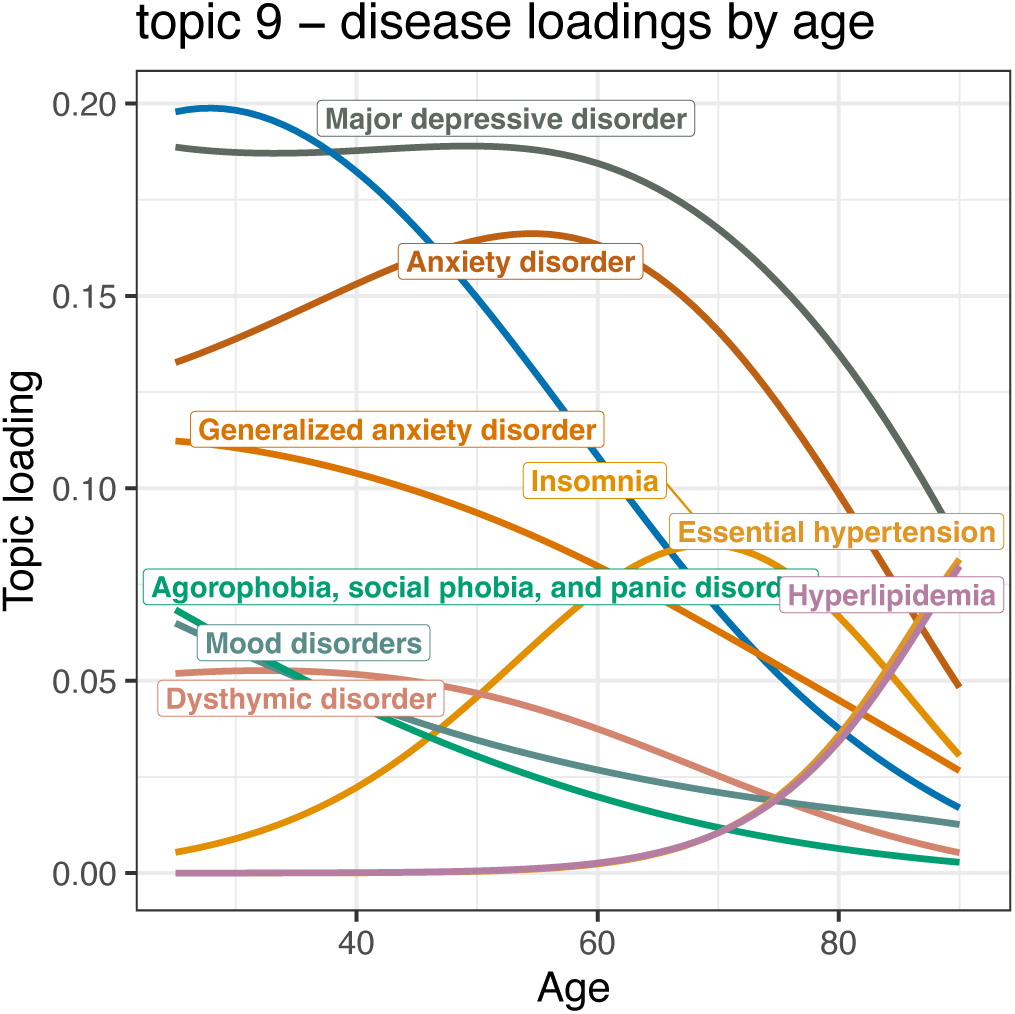
ATM Disease-Topic loadings: T09 PPM.

**Fig S10.**
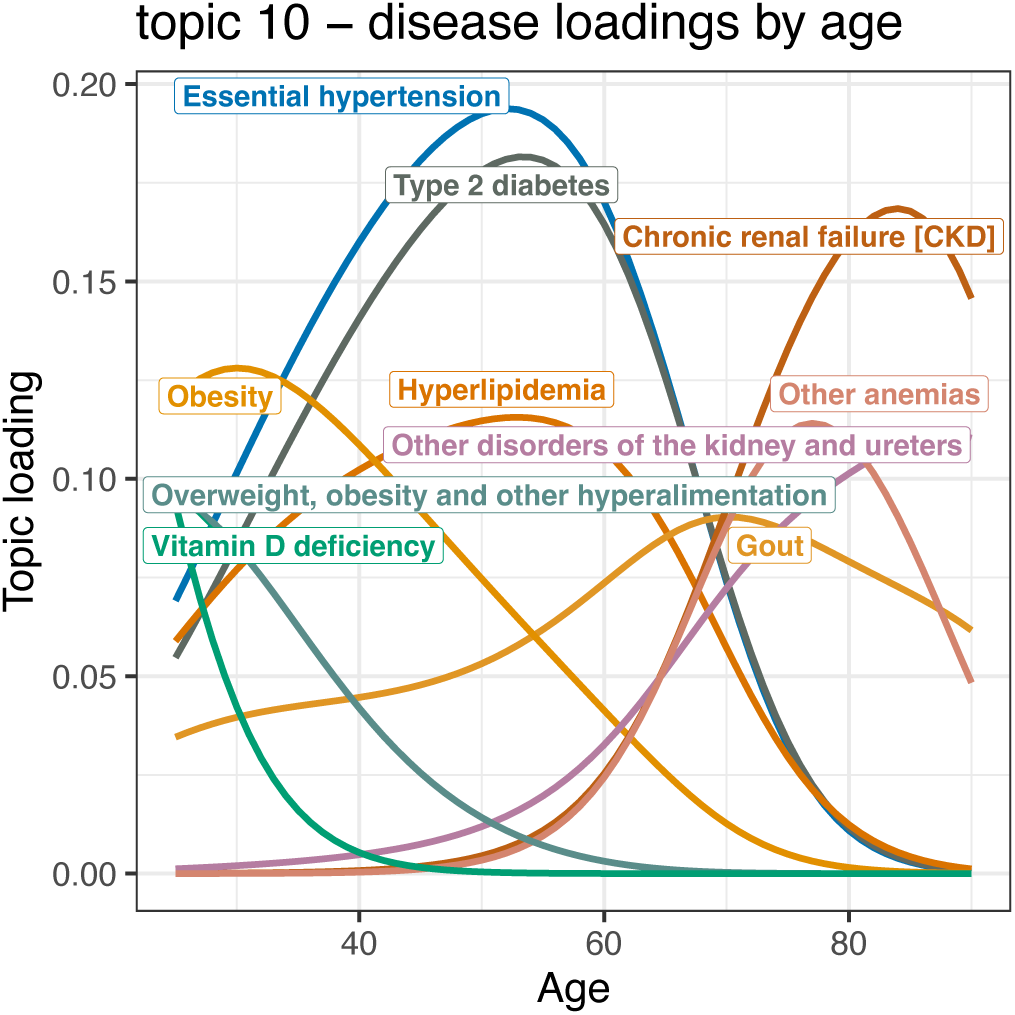
ATM Disease-Topic loadings: T10 BMK.

**Fig S11.**
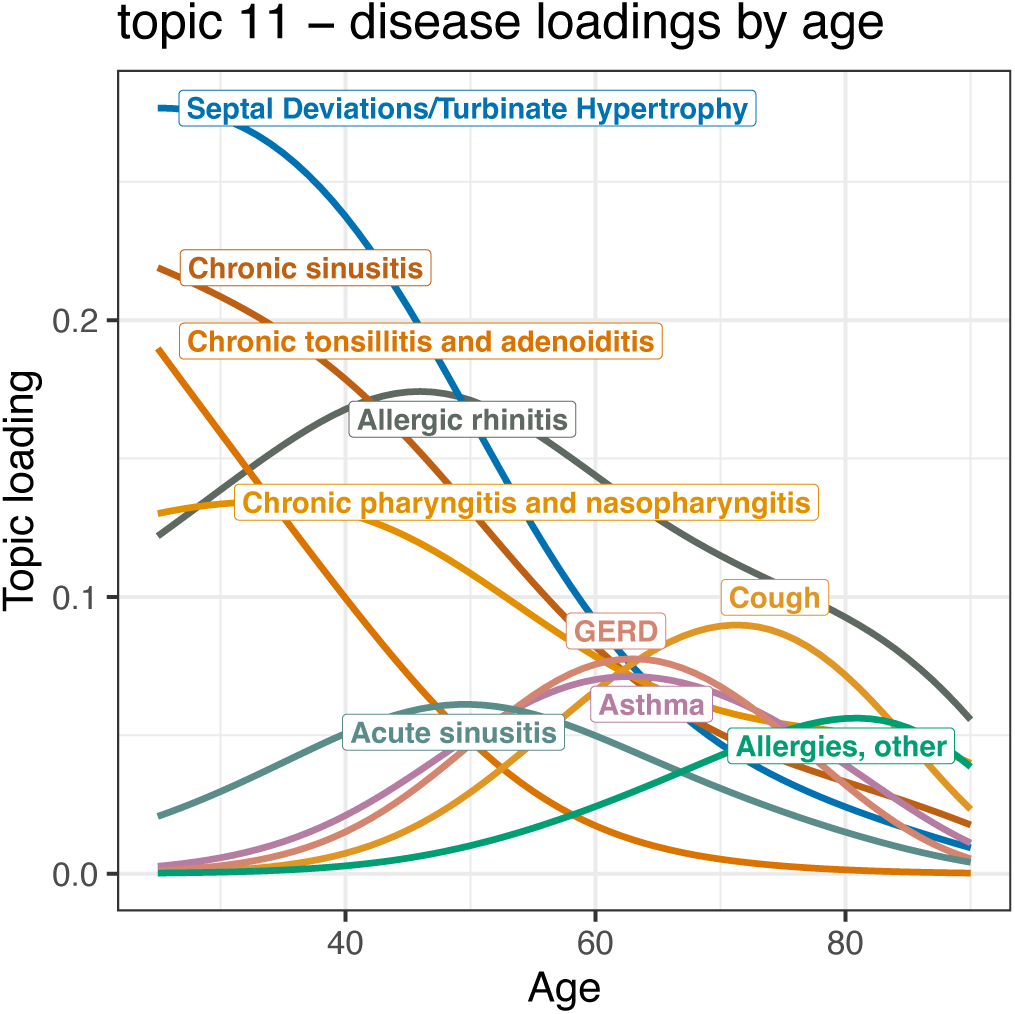
ATM Disease-Topic loadings: T11 IIL.

**Fig S12.**
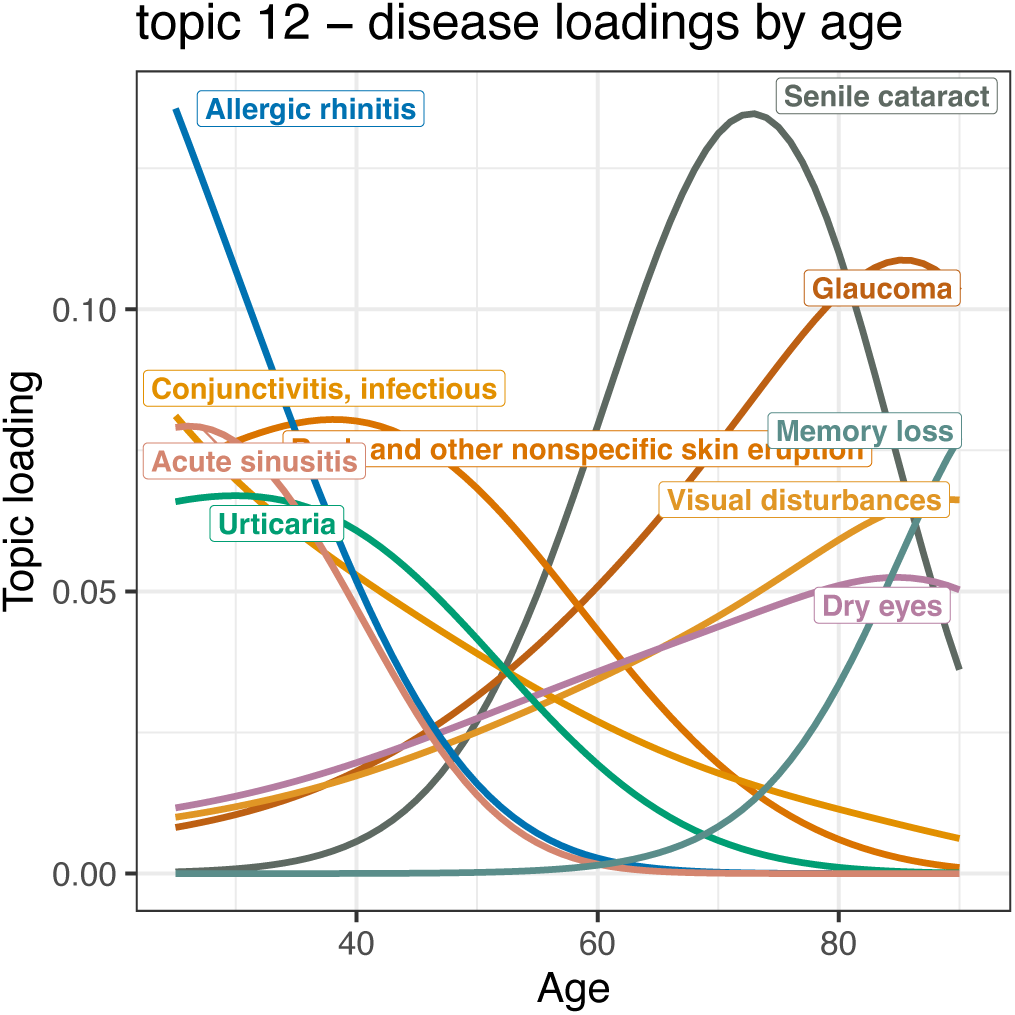
ATM Disease-Topic loadings: T12 IIC.

**Fig S13.**
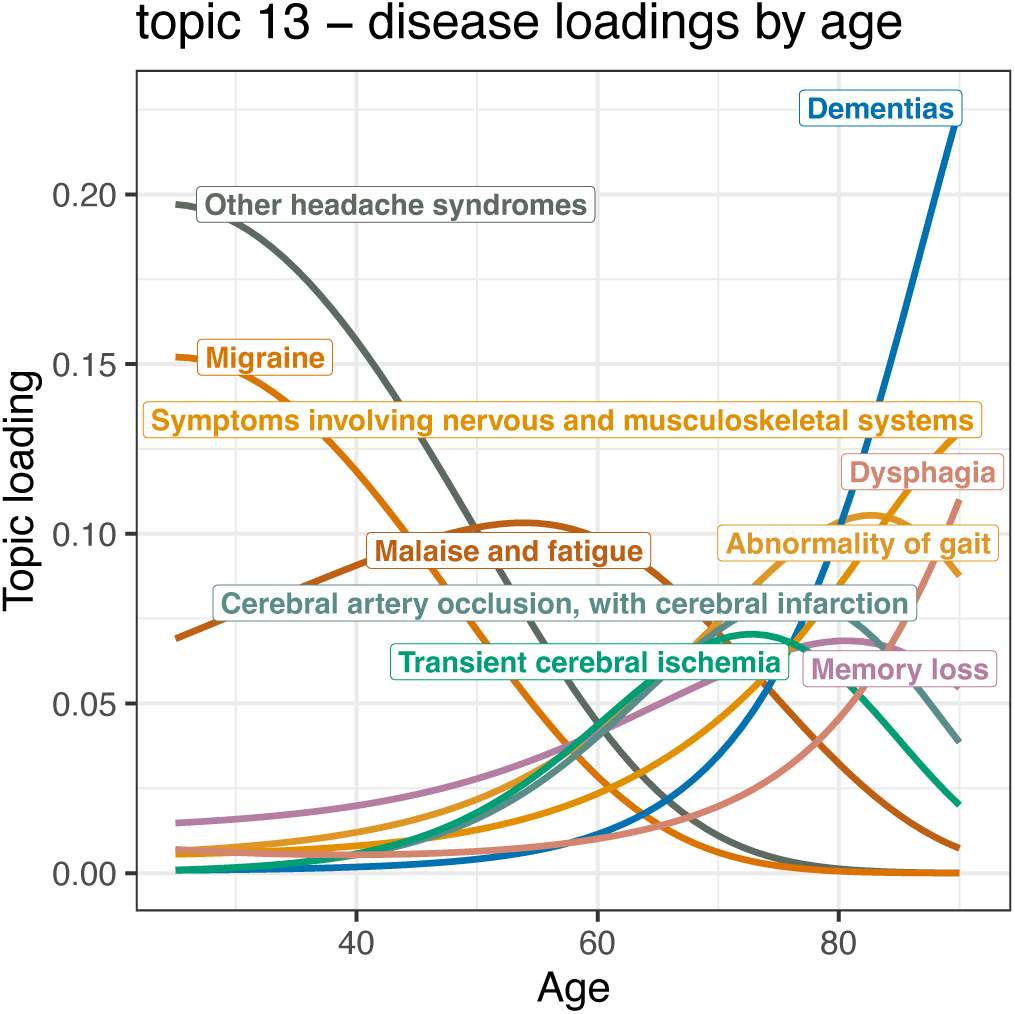
ATM Disease-Topic loadings: T13 PFV.

**Fig S14.**
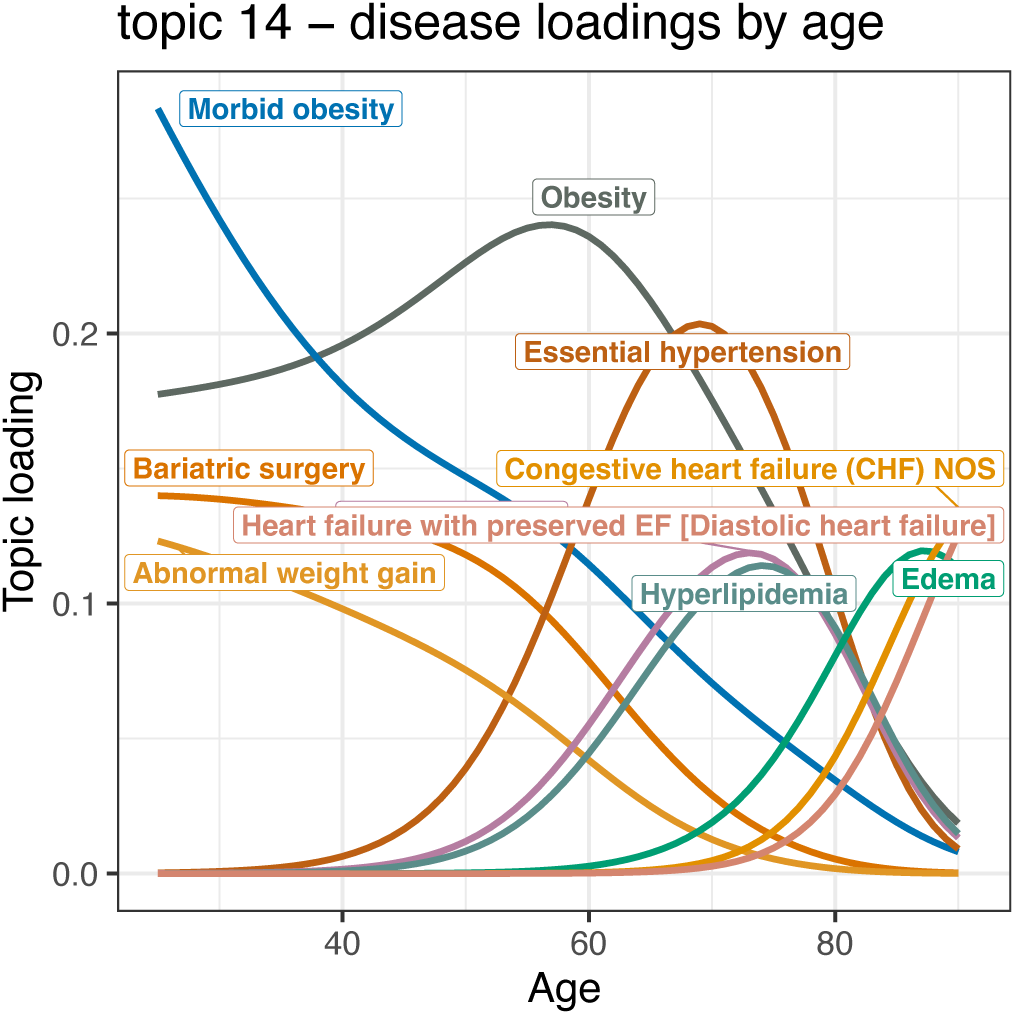
ATM Disease-Topic loadings: T14 BGM.

**Fig S15.**
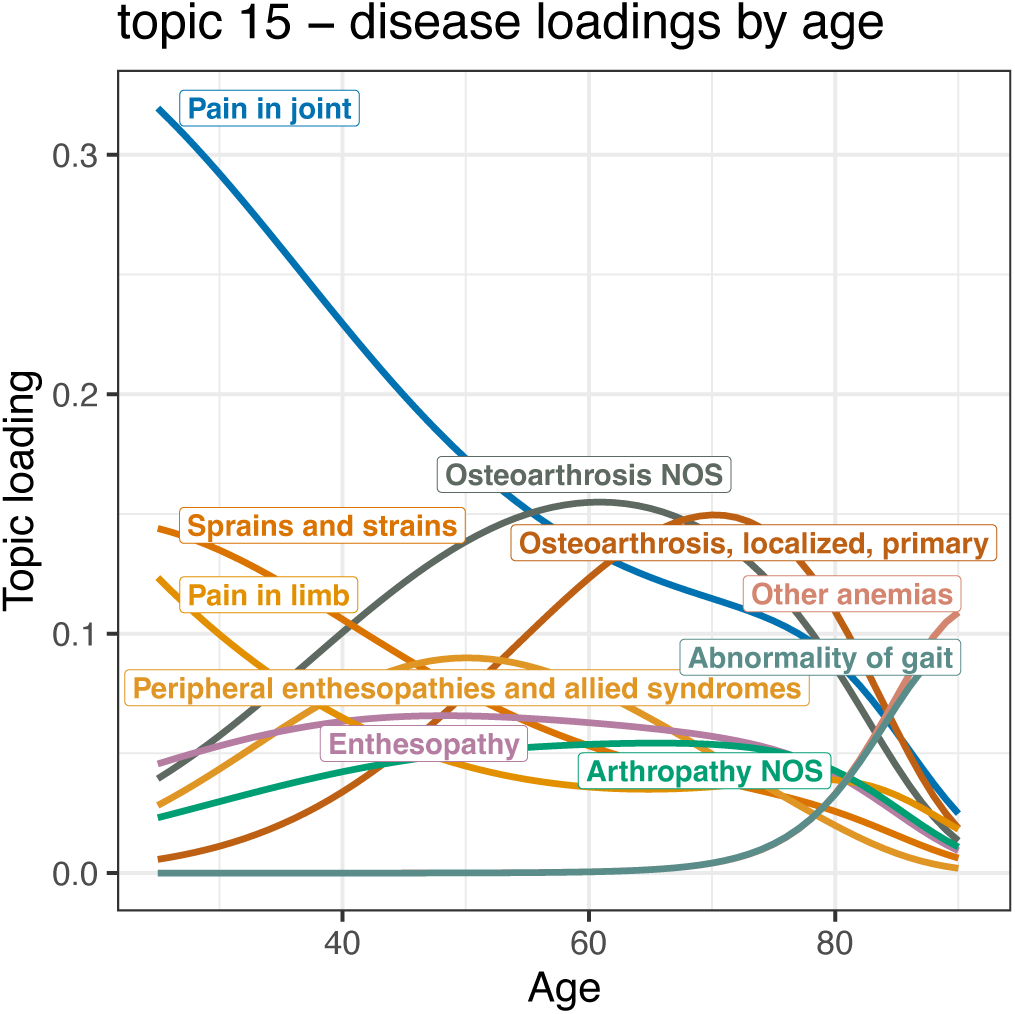
ATM Disease-Topic loadings: T15 OOO.

**Fig S16.**
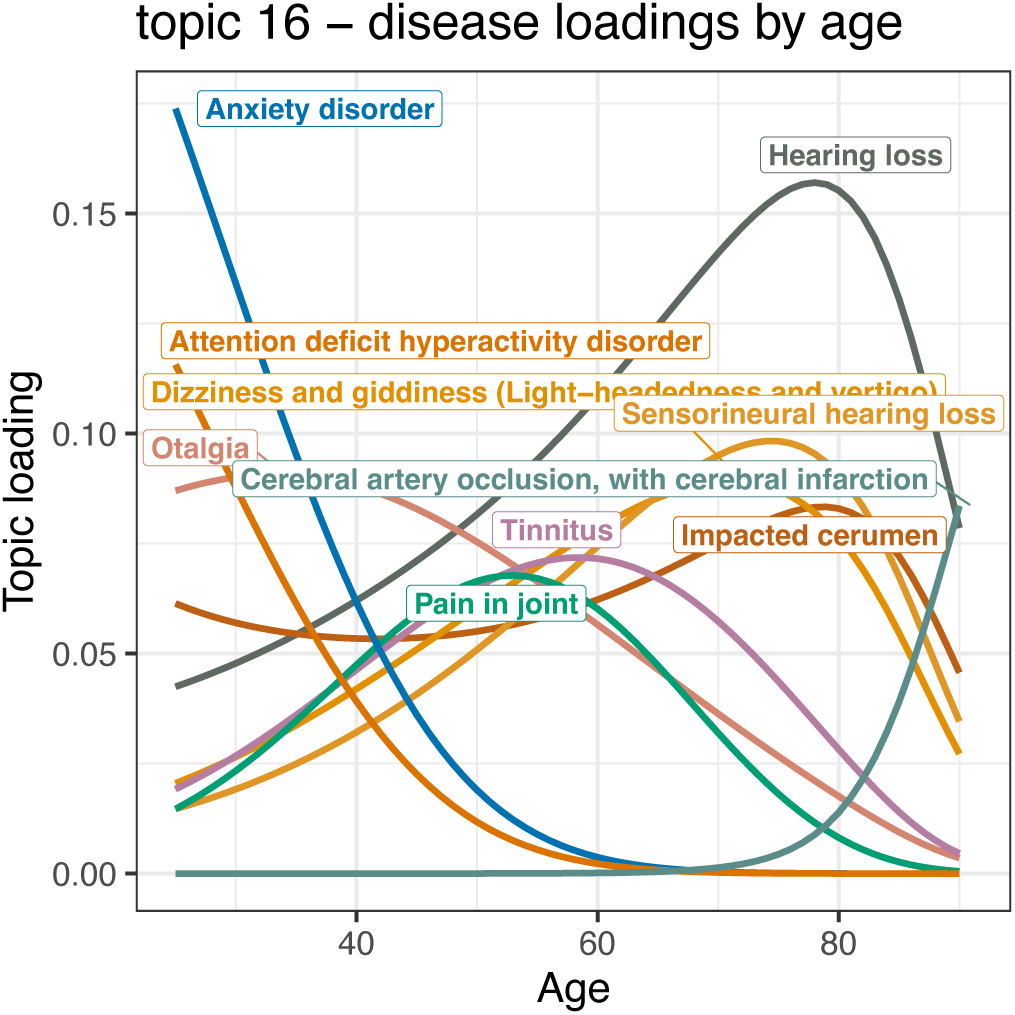
ATM Disease-Topic loadings: T16 PBM.

**Fig S17.**
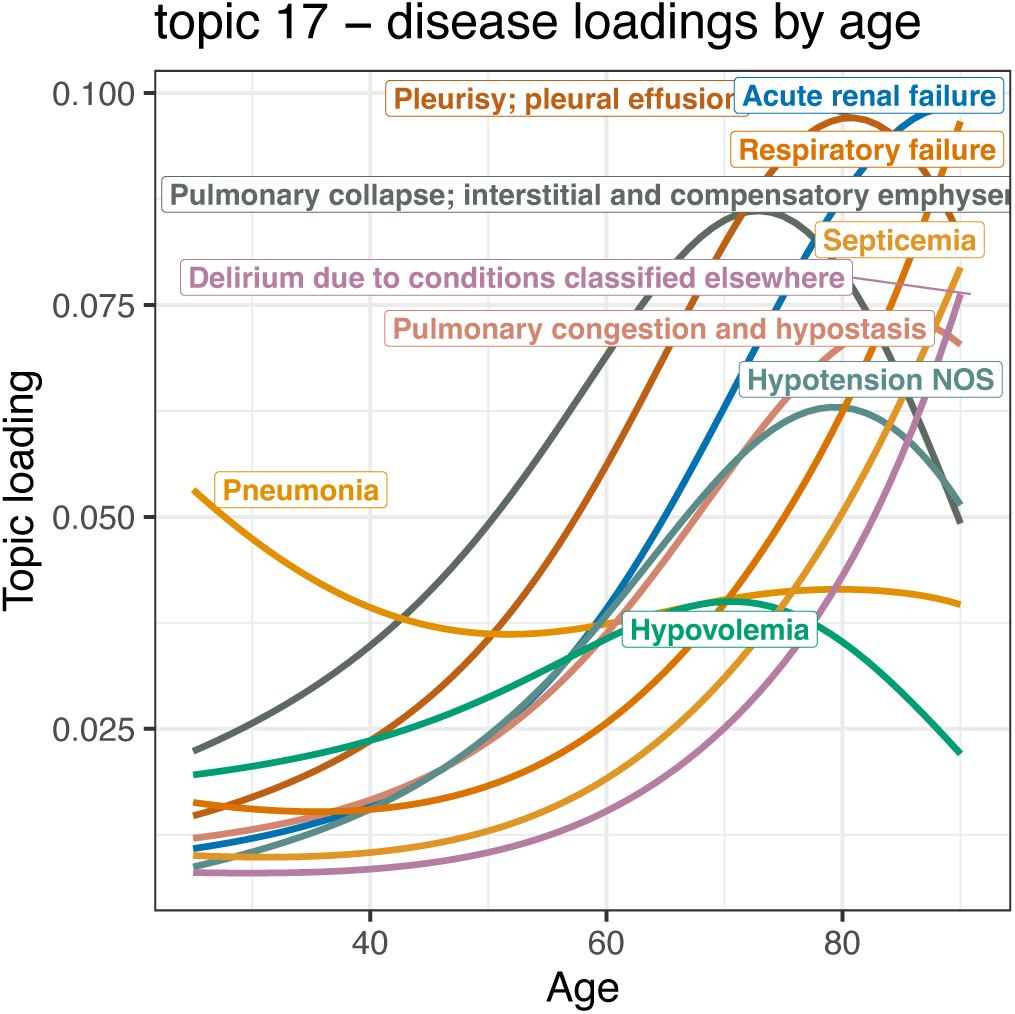
ATM Disease-Topic loadings: T17 VKL.

**Fig S18.**
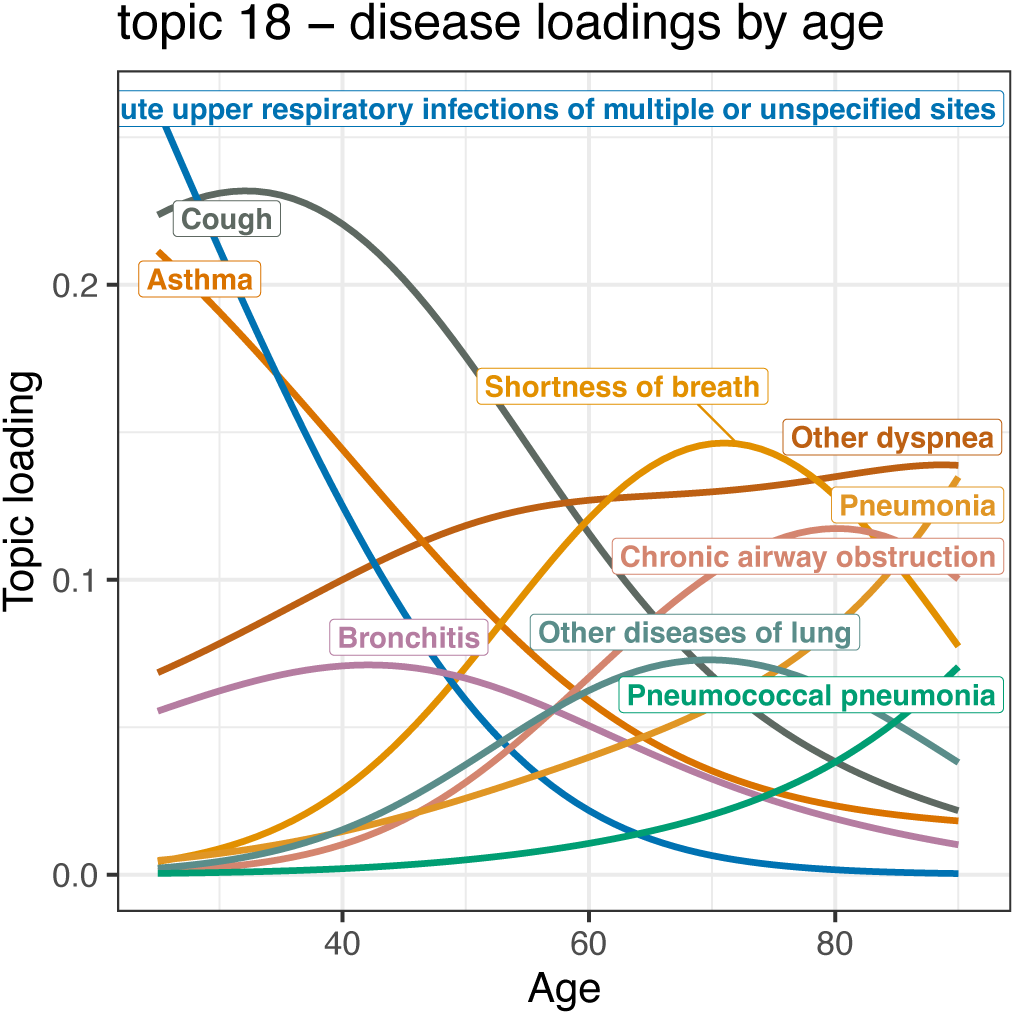
ATM Disease-Topic loadings: T18 ILL.

**Fig S19.**
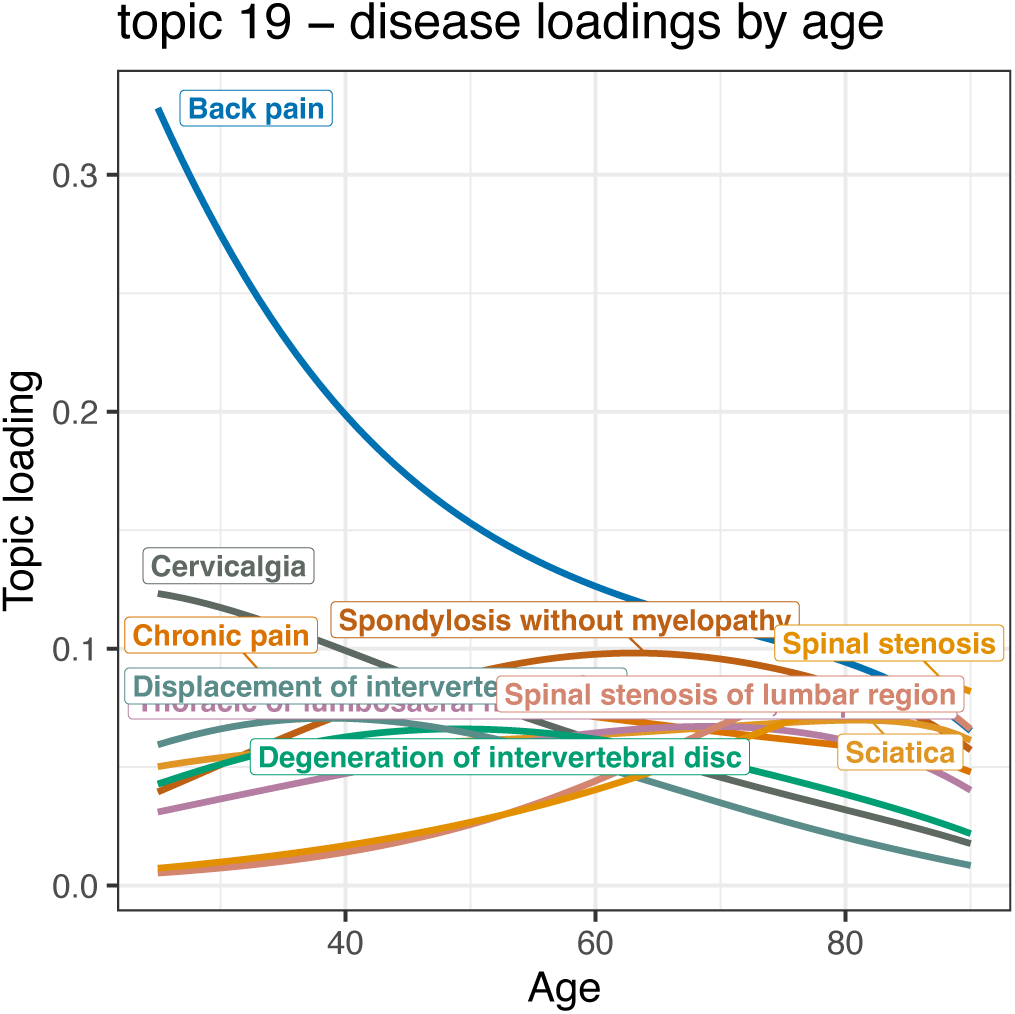
ATM Disease-Topic loadings: T19 SSS.

**Supplementary Table 1a:** Data distributions in combined cohort.

| Variable | Info | Overall | Spread | % Missing |
| --- | --- | --- | --- | --- |
| n |  | 156,441 |  |  |
| Age at first encounter | (median [IQR]) | 48.7 | [36.4, 59.3] | 0.3 |
| N encounters | (median [IQR]) | 72.0 | [28.0, 157.0] | 0.0 |
| Age | (median [IQR]) | 56.3 | [44.7, 66.6] | 0.1 |
| BMI | (median [IQR]) | 30.8 | [27.2, 35.0] | 0.1 |
| Population | (n (%)) |  |  | 0.0 |
| African |  | 10509 | (6.7) |  |
| Asian |  | 4172 | (2.7) |  |
| European |  | 129016 | (82.5) |  |
| Hispanic |  | 3230 | (2.1) |  |
| Other |  | 9514 | (6.1) |  |
| Sex (male) | (mean (SD)) | 0.582 | (0.493) | 0.0 |
| T01.IPK | (mean (SD)) | 0.063 | (0.112) | 4.7 |
| T02.UMU | (mean (SD)) | 0.050 | (0.092) | 4.7 |
| T03.IMG | (mean (SD)) | 0.050 | (0.073) | 4.7 |
| T04.IVO | (mean (SD)) | 0.051 | (0.089) | 4.7 |
| T05.HVK | (mean (SD)) | 0.050 | (0.104) | 4.7 |
| T06.BMV | (mean (SD)) | 0.051 | (0.090) | 4.7 |
| T07.GGG | (mean (SD)) | 0.048 | (0.079) | 4.7 |
| T08.DDD | (mean (SD)) | 0.057 | (0.123) | 4.7 |
| T09.PPM | (mean (SD)) | 0.048 | (0.097) | 4.7 |
| T10.BMK | (mean (SD)) | 0.048 | (0.078) | 4.7 |
| T11.IIL | (mean (SD)) | 0.040 | (0.087) | 4.7 |
| T12.IIC | (mean (SD)) | 0.054 | (0.089) | 4.7 |
| T13.PFV | (mean (SD)) | 0.053 | (0.094) | 4.7 |
| T14.BGM | (mean (SD)) | 0.051 | (0.090) | 4.7 |
| T15.OOO | (mean (SD)) | 0.072 | (0.135) | 4.7 |
| T16.PBM | (mean (SD)) | 0.047 | (0.083) | 4.7 |
| T17.VKL | (mean (SD)) | 0.058 | (0.110) | 4.7 |
| T18.ILL | (mean (SD)) | 0.049 | (0.086) | 4.7 |
| T19.SSS | (mean (SD)) | 0.059 | (0.118) | 4.7 |
| AHI | (median [IQR]) | 17.2 | [10.1, 28.6] | 95.8 |
| T90 | (median [IQR]) | 0.1 | [0.0, 1.4] | 95.8 |
| ArBdn | (median [IQR]) | 126.8 | [94.1, 168.8] | 95.8 |
| ArThr | (median [IQR]) | 122.6 | [111.8, 136.3] | 95.8 |
| ChxDly | (median [IQR]) | 12.5 | [10.8, 14.8] | 95.8 |
| HypBdn | (median [IQR]) | 17.4 | [8.0, 36.5] | 95.8 |
| HRBdn | (median [IQR]) | 131.3 | [72.3, 234.1] | 95.8 |
| LG1 | (median [IQR]) | 0.5 | [0.4, 0.6] | 95.8 |
| LGN | (median [IQR]) | 0.4 | [0.3, 0.5] | 95.8 |
| VntBdn | (median [IQR]) | 262.2 | [136.7, 493.2] | 95.8 |
| Vpassive | (median [IQR]) | 96.0 | [93.3, 97.8] | 95.8 |
| Vactive | (median [IQR]) | 101.5 | [97.8, 103.8] | 95.8 |
| Vcomp | (median [IQR]) | 4.6 | [3.2, 6.6] | 95.8 |

**Supplementary Table 1b:**
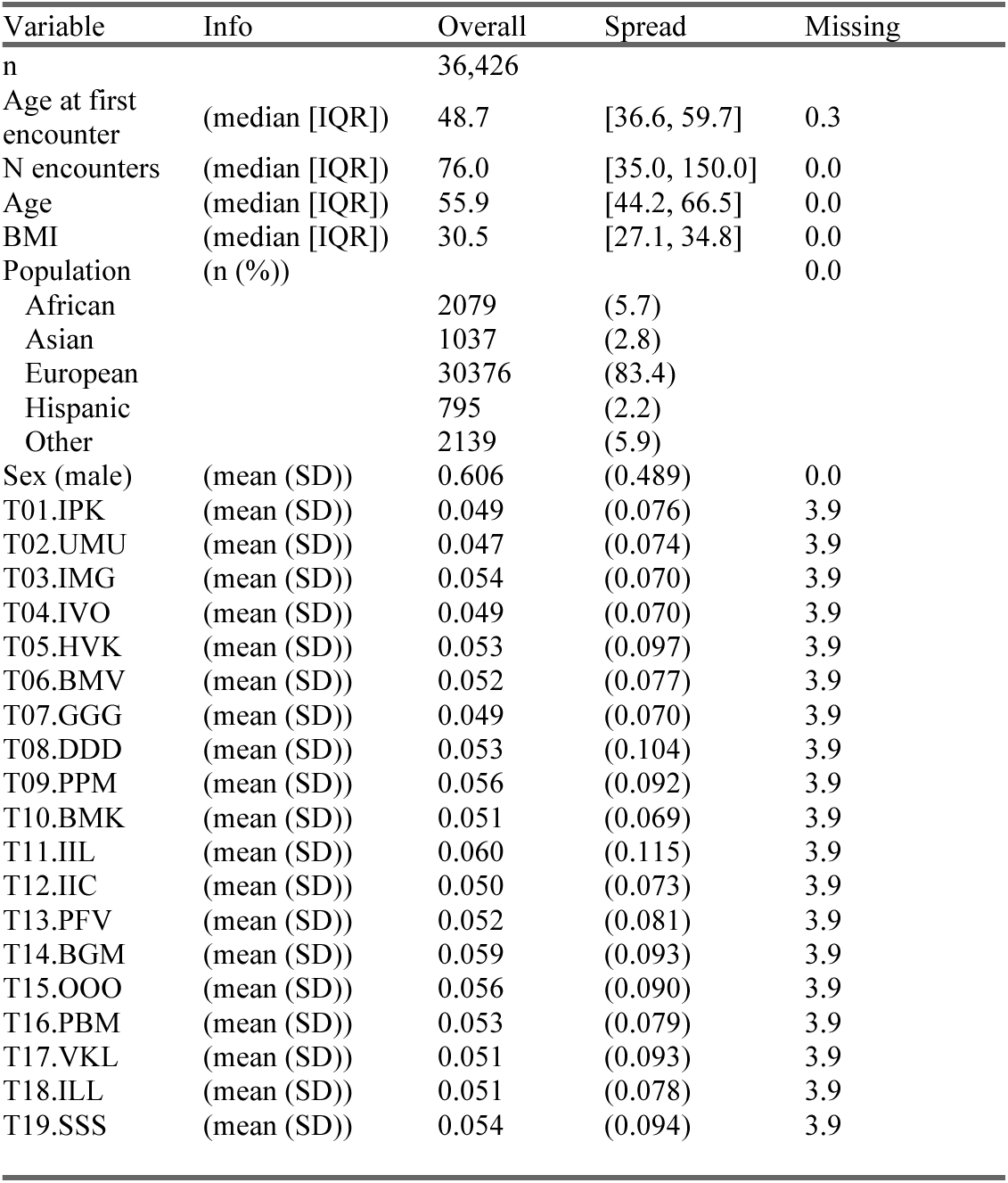
Distributions in Case-only training cohort.

**Supplementary Table 1c:** Distributions in testing cohorts, by cohort.

| Variable |  | Faulkner clinical<br>PSG cohort |  | RPDR OSA<br>case-control |  |
| --- | --- | --- | --- | --- | --- |
| n |  | 6220 |  | 34384 |  |
| Age at first encounter | (median [IQR]) | 41.0 | [29.8, 52.6] | 47.5 | [36.5, 57.4] |
| N encounters | (median [IQR]) | 144.0 | [66.0, 271.0] | 101.0 | [39.0, 204.0] |
| Age | (median [IQR]) | 55.2 | [41.0, 67.0] | 57.7 | [47.3, 67.1] |
| BMI | (median [IQR]) | 29.2 | [25.5, 34.6] | 30.5 | [27.1, 34.9] |
| Population (%) |  |  |  |  |  |
| African |  | 832 | (13.4) | 2515 | ( 7.3) |
| Asian |  | 223 | ( 3.6) | 897 | ( 2.6) |
| European |  | 4010 | (64.5) | 27861 | (81.0) |
| Hispanic |  | 519 | ( 8.3) | 885 | ( 2.6) |
| Other |  | 636 | (10.2) | 2226 | ( 6.5) |
| Sex (male) | (mean (SD)) | 0.381 | (0.486) | 0.548 | (0.498) |
| T01.IPK | (mean (SD)) | 0.051 | (0.068) | 0.065 | (0.111) |
| T02.UMU | (mean (SD)) | 0.052 | (0.074) | 0.051 | (0.092) |
| T03.IMG | (mean (SD)) | 0.047 | (0.061) | 0.050 | (0.072) |
| T04.IVO | (mean (SD)) | 0.054 | (0.068) | 0.054 | (0.089) |
| T05.HVK | (mean (SD)) | 0.050 | (0.090) | 0.050 | (0.103) |
| T06.BMV | (mean (SD)) | 0.040 | (0.061) | 0.048 | (0.086) |
| T07.GGG | (mean (SD)) | 0.050 | (0.069) | 0.047 | (0.077) |
| T08.DDD | (mean (SD)) | 0.064 | (0.111) | 0.062 | (0.125) |
| T09.PPM | (mean (SD)) | 0.064 | (0.093) | 0.048 | (0.096) |
| T10.BMK | (mean (SD)) | 0.037 | (0.055) | 0.047 | (0.076) |
| T11.IIL | (mean (SD)) | 0.036 | (0.072) | 0.036 | (0.073) |
| T12.IIC | (mean (SD)) | 0.059 | (0.073) | 0.057 | (0.087) |
| T13.PFV | (mean (SD)) | 0.075 | (0.099) | 0.055 | (0.093) |
| T14.BGM | (mean (SD)) | 0.054 | (0.084) | 0.047 | (0.084) |
| T15.OOO | (mean (SD)) | 0.060 | (0.090) | 0.072 | (0.130) |
| T16.PBM | (mean (SD)) | 0.054 | (0.074) | 0.044 | (0.075) |
| T17.VKL | (mean (SD)) | 0.049 | (0.083) | 0.061 | (0.111) |
| T18.ILL | (mean (SD)) | 0.044 | (0.067) | 0.048 | (0.082) |
| T19.SSS | (mean (SD)) | 0.060 | (0.092) | 0.058 | (0.113) |
| AHI | (median [IQR]) | 17.5 | [10.2, 29.0] | NA | [NA, NA] |
| T90 | (median [IQR]) | 0.1 | [0.0, 1.6] | NA | [NA, NA] |
| ArBdn | (median [IQR]) | 126.4 | [94.0, 168.6] | NA | [NA, NA] |
| ArThr | (median [IQR]) | 123.1 | [112.3, 136.7] | NA | [NA, NA] |
| ChxDly | (median [IQR]) | 12.5 | [10.8, 14.7] | NA | [NA, NA] |
| HypBdn | (median [IQR]) | 18.1 | [8.4, 37.1] | NA | [NA, NA] |
| HRBdn | (median [IQR]) | 132.3 | [72.2, 236.6] | NA | [NA, NA] |
| LG1 | (median [IQR]) | 0.5 | [0.4, 0.6] | NA | [NA, NA] |
| LGN | (median [IQR]) | 0.4 | [0.3, 0.5] | NA | [NA, NA] |
| VntBdn | (median [IQR]) | 269.7 | [140.4, 499.6] | NA | [NA, NA] |
| Vpassive | (median [IQR]) | 96.0 | [93.3, 97.8] | NA | [NA, NA] |
| Vactive | (median [IQR]) | 101.5 | [97.8, 103.8] | NA | [NA, NA] |
| Vcomp | (median [IQR]) | 4.6 | [3.2, 6.7] | NA | [NA, NA] |

**Supplementary Table 2:**
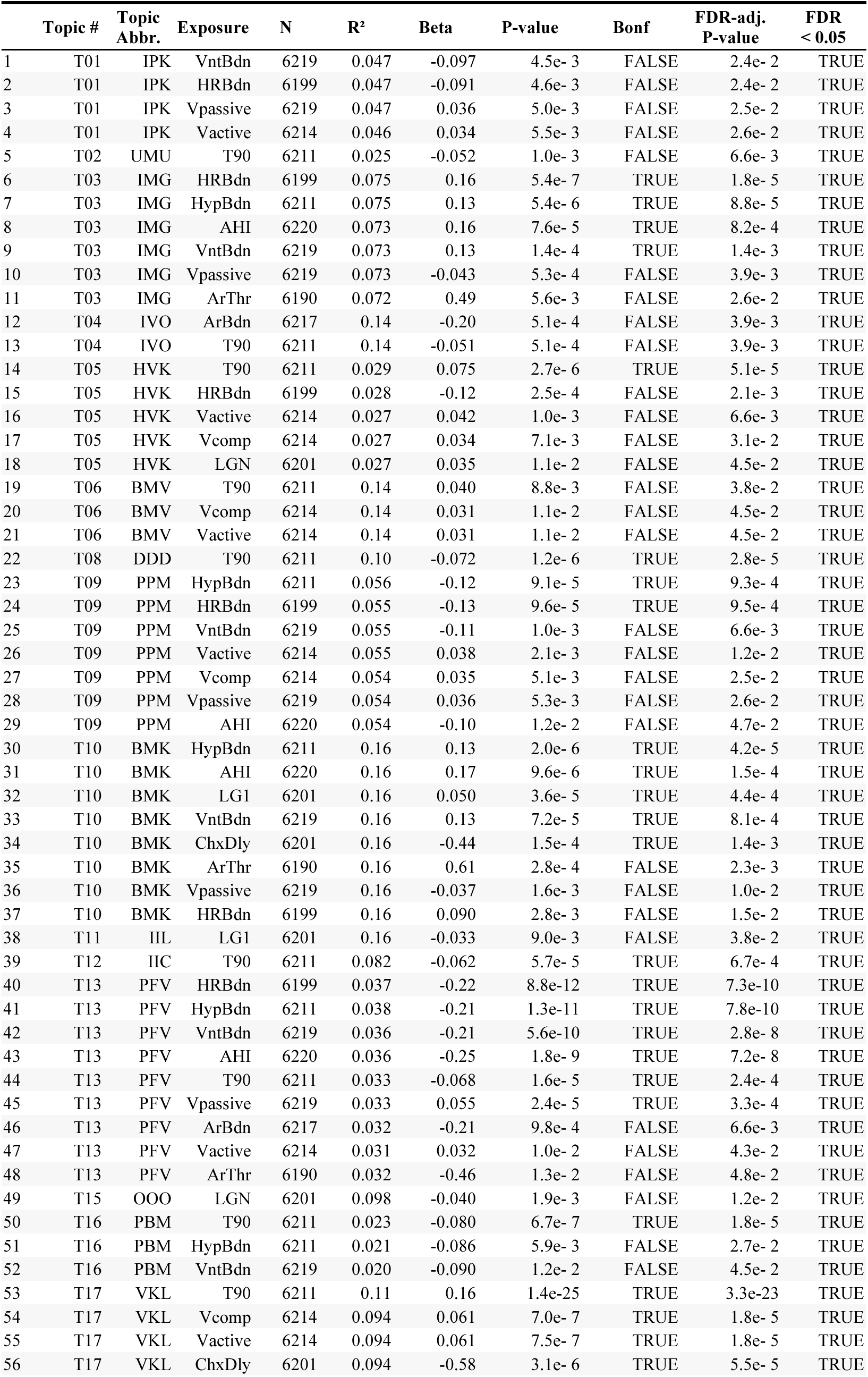
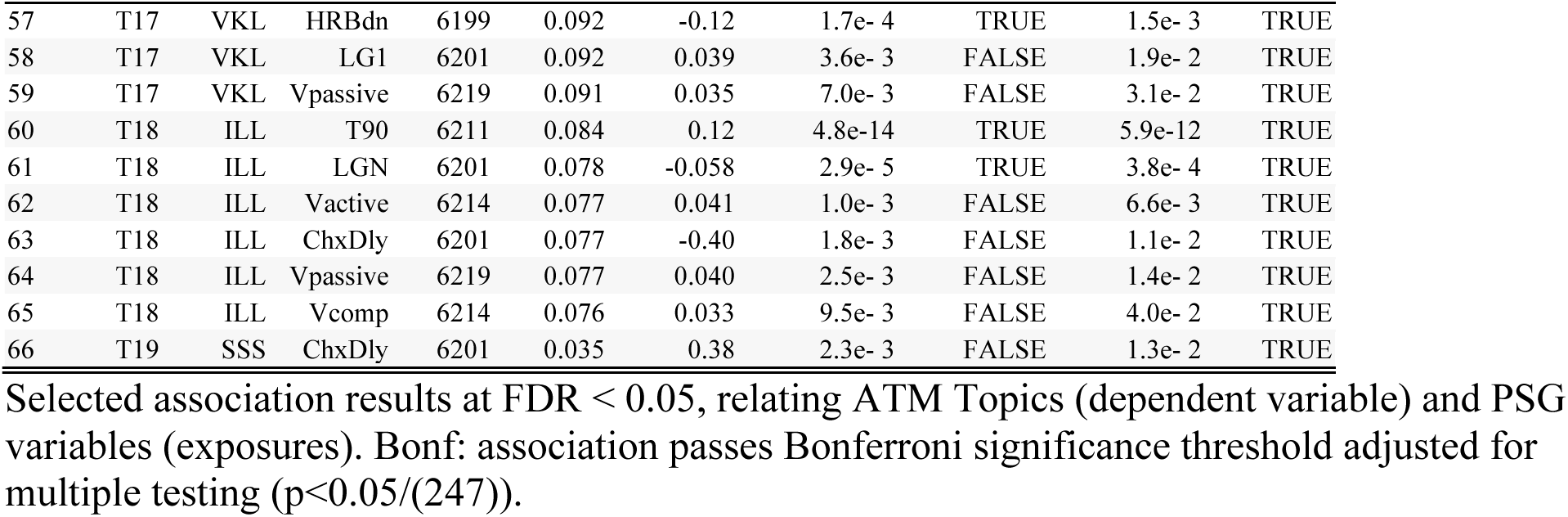
Associations between ATM topics and PSG measures.

**Supplementary Table 3.**
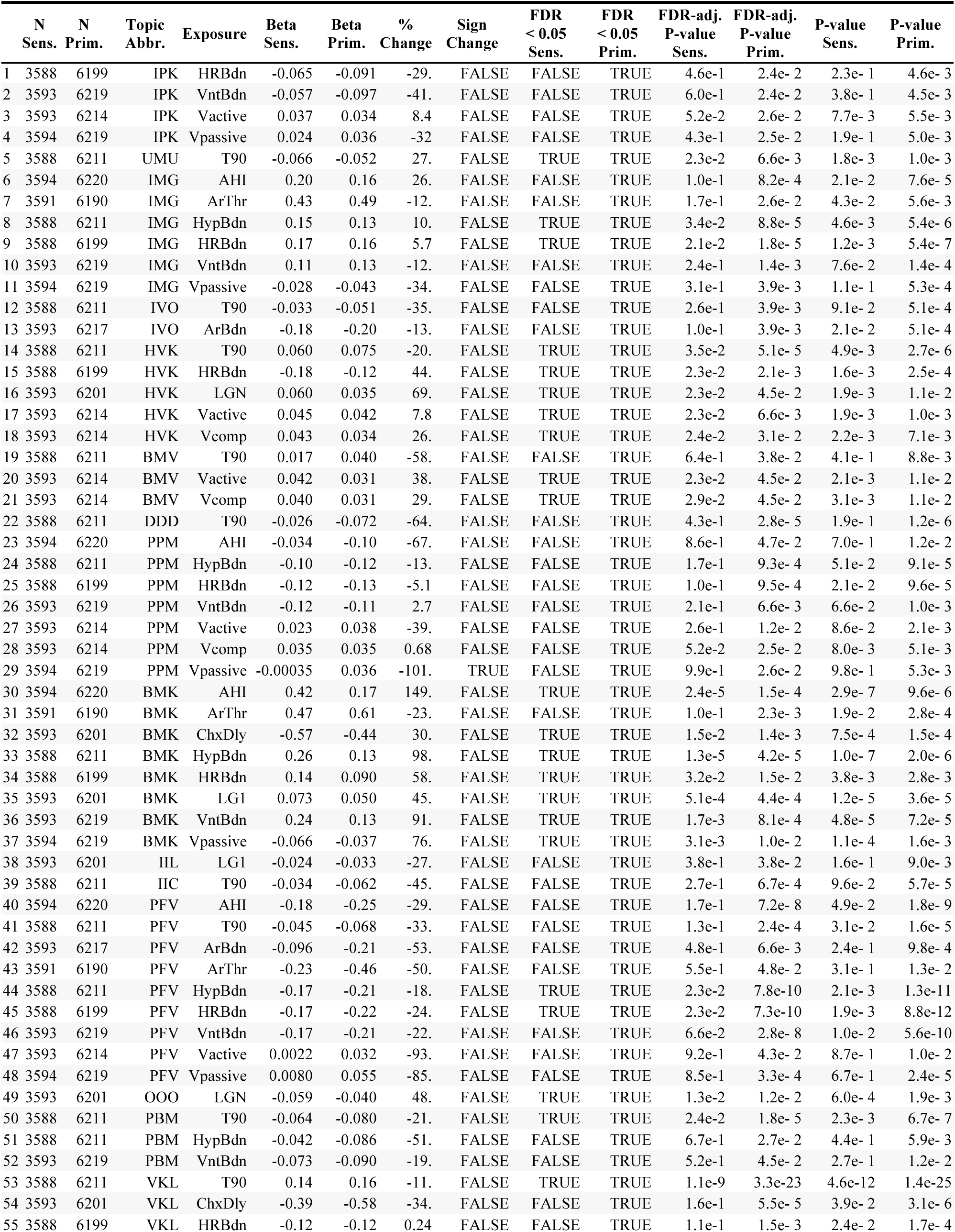
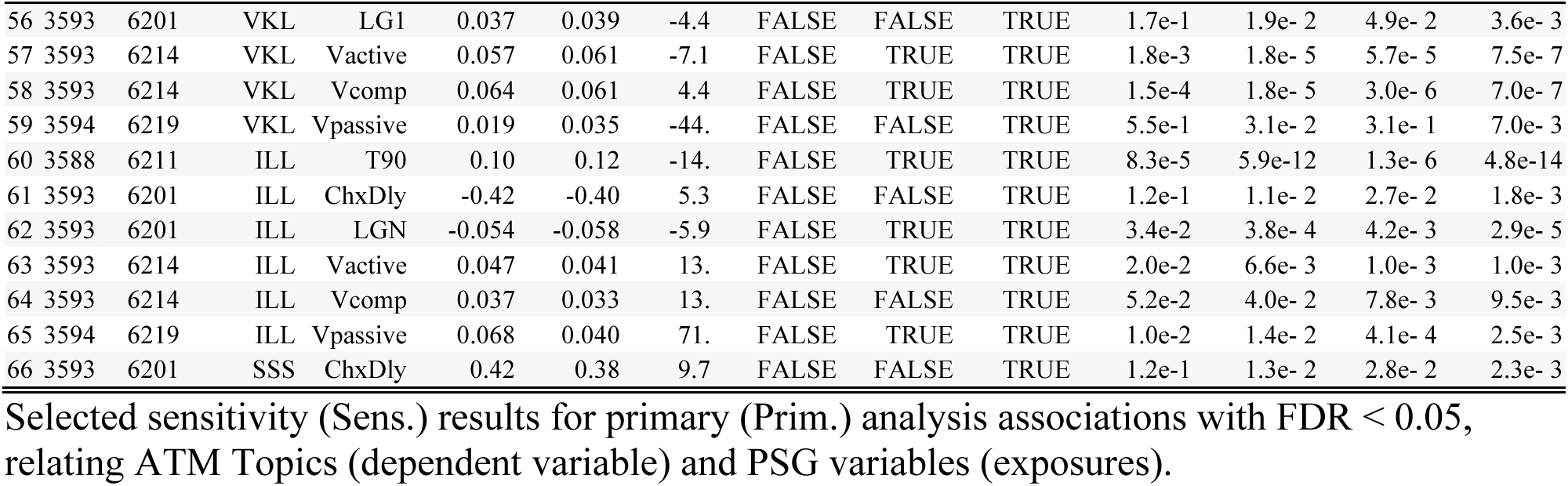
Sensitivity Analysis: OSA subset (AHI ≥ 15)

|  | N | N | Topic | Exposure | Beta | Beta | % | Sign | FDR | FDR | FDR-adj. | FDR-adj. | P-value | P-value |
| --- | --- | --- | --- | --- | --- | --- | --- | --- | --- | --- | --- | --- | --- | --- |
|  | Sens. | Prim. | Abbr. |  | Sens. | Prim. | Change | Change | < 0.05 | < 0.05 | P-value | P-value | Sens. | Prim. |
|  |  |  |  |  |  |  |  |  | Sens. | Prim. | Sens. | Prim. |  |  |
| 1 | 3588 | 6199 | IPK | HRBdn | -0.065 | -0.091 | -29. | FALSE | FALSE | TRUE | 4.6e-1 | 2.4e-2 | 2.3e-1 | 4.6e-3 |
| 2 | 3593 | 6219 | IPK | VntBdn | -0.057 | -0.097 | -41. | FALSE | FALSE | TRUE | 6.0e-1 | 2.4e-2 | 3.8e-1 | 4.5e-3 |
| 3 | 3593 | 6214 | IPK | Vactive | 0.037 | 0.034 | 8.4 | FALSE | FALSE | TRUE | 5.2e-2 | 2.6e-2 | 7.7e-3 | 5.5e-3 |
| 4 | 3594 | 6219 | IPK | Vpassive | 0.024 | 0.036 | -32 | FALSE | FALSE | TRUE | 4.3e-1 | 2.5e-2 | 1.9e-1 | 5.0e-3 |
| 5 | 3588 | 6211 | UMU | T90 | -0.066 | -0.052 | 27. | FALSE | TRUE | TRUE | 2.3e-2 | 6.6e-3 | 1.8e-3 | 1.0e-3 |
| 6 | 3594 | 6220 | IMG | AHI | 0.20 | 0.16 | 26. | FALSE | FALSE | TRUE | 1.0e-1 | 8.2e-4 | 2.1e-2 | 7.6e-5 |
| 7 | 3591 | 6190 | IMG | ArThr | 0.43 | 0.49 | -12. | FALSE | FALSE | TRUE | 1.7e-1 | 2.6e-2 | 4.3e-2 | 5.6e-3 |
| 8 | 3588 | 6211 | IMG | HypBdn | 0.15 | 0.13 | 10. | FALSE | TRUE | TRUE | 3.4e-2 | 8.8e-5 | 4.6e-3 | 5.4e-6 |
| 9 | 3588 | 6199 | IMG | HRBdn | 0.17 | 0.16 | 5.7 | FALSE | TRUE | TRUE | 2.1e-2 | 1.8e-5 | 1.2e-3 | 5.4e-7 |
| 10 | 3593 | 6219 | IMG | VntBdn | 0.11 | 0.13 | -12. | FALSE | FALSE | TRUE | 2.4e-1 | 1.4e-3 | 7.6e-2 | 1.4e-4 |
| 11 | 3594 | 6219 | IMG | Vpassive | -0.028 | -0.043 | -34. | FALSE | FALSE | TRUE | 3.1e-1 | 3.9e-3 | 1.1e-1 | 5.3e-4 |
| 12 | 3588 | 6211 | IVO | T90 | -0.033 | -0.051 | -35. | FALSE | FALSE | TRUE | 2.6e-1 | 3.9e-3 | 9.1e-2 | 5.1e-4 |
| 13 | 3593 | 6217 | IVO | ArBdn | -0.18 | -0.20 | -13. | FALSE | FALSE | TRUE | 1.0e-1 | 3.9e-3 | 2.1e-2 | 5.1e-4 |
| 14 | 3588 | 6211 | HVK | T90 | 0.060 | 0.075 | -20. | FALSE | TRUE | TRUE | 3.5e-2 | 5.1e-5 | 4.9e-3 | 2.7e-6 |
| 15 | 3588 | 6199 | HVK | HRBdn | -0.18 | -0.12 | 44. | FALSE | TRUE | TRUE | 2.3e-2 | 2.1e-3 | 1.6e-3 | 2.5e-4 |
| 16 | 3593 | 6201 | HVK | LGN | 0.060 | 0.035 | 69. | FALSE | TRUE | TRUE | 2.3e-2 | 4.5e-2 | 1.9e-3 | 1.1e-2 |
| 17 | 3593 | 6214 | HVK | Vactive | 0.045 | 0.042 | 7.8 | FALSE | TRUE | TRUE | 2.3e-2 | 6.6e-3 | 1.9e-3 | 1.0e-3 |
| 18 | 3593 | 6214 | HVK | Vcomp | 0.043 | 0.034 | 26. | FALSE | TRUE | TRUE | 2.4e-2 | 3.1e-2 | 2.2e-3 | 7.1e-3 |
| 19 | 3588 | 6211 | BMV | T90 | 0.017 | 0.040 | -58. | FALSE | FALSE | TRUE | 6.4e-1 | 3.8e-2 | 4.1e-1 | 8.8e-3 |
| 20 | 3593 | 6214 | BMV | Vactive | 0.042 | 0.031 | 38. | FALSE | TRUE | TRUE | 2.3e-2 | 4.5e-2 | 2.1e-3 | 1.1e-2 |
| 21 | 3593 | 6214 | BMV | Vcomp | 0.040 | 0.031 | 29. | FALSE | TRUE | TRUE | 2.9e-2 | 4.5e-2 | 3.1e-3 | 1.1e-2 |
| 22 | 3588 | 6211 | DDD | T90 | -0.026 | -0.072 | -64. | FALSE | FALSE | TRUE | 4.3e-1 | 2.8e-5 | 1.9e-1 | 1.2e-6 |
| 23 | 3594 | 6220 | PPM | AHI | -0.034 | -0.10 | -67. | FALSE | FALSE | TRUE | 8.6e-1 | 4.7e-2 | 7.0e-1 | 1.2e-2 |
| 24 | 3588 | 6211 | PPM | HypBdn | -0.10 | -0.12 | -13. | FALSE | FALSE | TRUE | 1.7e-1 | 9.3e-4 | 5.1e-2 | 9.1e-5 |
| 25 | 3588 | 6199 | PPM | HRBdn | -0.12 | -0.13 | -5.1 | FALSE | FALSE | TRUE | 1.0e-1 | 9.5e-4 | 2.1e-2 | 9.6e-5 |
| 26 | 3593 | 6219 | PPM | VntBdn | -0.12 | -0.11 | 2.7 | FALSE | FALSE | TRUE | 2.1e-1 | 6.6e-3 | 6.6e-2 | 1.0e-3 |
| 27 | 3593 | 6214 | PPM | Vactive | 0.023 | 0.038 | -39. | FALSE | FALSE | TRUE | 2.6e-1 | 1.2e-2 | 8.6e-2 | 2.1e-3 |
| 28 | 3593 | 6214 | PPM | Vcomp | 0.035 | 0.035 | 0.68 | FALSE | FALSE | TRUE | 5.2e-2 | 2.5e-2 | 8.0e-3 | 5.1e-3 |
| 29 | 3594 | 6219 | PPM | Vpassive | -0.00035 | 0.036 | -101. | TRUE | FALSE | TRUE | 9.9e-1 | 2.6e-2 | 9.8e-1 | 5.3e-3 |
| 30 | 3594 | 6220 | BMK | AHI | 0.42 | 0.17 | 149. | FALSE | TRUE | TRUE | 2.4e-5 | 1.5e-4 | 2.9e-7 | 9.6e-6 |
| 31 | 3591 | 6190 | BMK | ArThr | 0.47 | 0.61 | -23. | FALSE | FALSE | TRUE | 1.0e-1 | 2.3e-3 | 1.9e-2 | 2.8e-4 |
| 32 | 3593 | 6201 | BMK | ChxDly | -0.57 | -0.44 | 30. | FALSE | TRUE | TRUE | 1.5e-2 | 1.4e-3 | 7.5e-4 | 1.5e-4 |
| 33 | 3588 | 6211 | BMK | HypBdn | 0.26 | 0.13 | 98. | FALSE | TRUE | TRUE | 1.3e-5 | 4.2e-5 | 1.0e-7 | 2.0e-6 |
| 34 | 3588 | 6199 | BMK | HRBdn | 0.14 | 0.090 | 58. | FALSE | TRUE | TRUE | 3.2e-2 | 1.5e-2 | 3.8e-3 | 2.8e-3 |
| 35 | 3593 | 6201 | BMK | LG1 | 0.073 | 0.050 | 45. | FALSE | TRUE | TRUE | 5.1e-4 | 4.4e-4 | 1.2e-5 | 3.6e-5 |
| 36 | 3593 | 6219 | BMK | VntBdn | 0.24 | 0.13 | 91. | FALSE | TRUE | TRUE | 1.7e-3 | 8.1e-4 | 4.8e-5 | 7.2e-5 |
| 37 | 3594 | 6219 | BMK | Vpassive | -0.066 | -0.037 | 76. | FALSE | TRUE | TRUE | 3.1e-3 | 1.0e-2 | 1.1e-4 | 1.6e-3 |
| 38 | 3593 | 6201 | IIL | LG1 | -0.024 | -0.033 | -27. | FALSE | FALSE | TRUE | 3.8e-1 | 3.8e-2 | 1.6e-1 | 9.0e-3 |
| 39 | 3588 | 6211 | IIC | T90 | -0.034 | -0.062 | -45. | FALSE | FALSE | TRUE | 2.7e-1 | 6.7e-4 | 9.6e-2 | 5.7e-5 |
| 40 | 3594 | 6220 | PFV | AHI | -0.18 | -0.25 | -29. | FALSE | FALSE | TRUE | 1.7e-1 | 7.2e-8 | 4.9e-2 | 1.8e-9 |
| 41 | 3588 | 6211 | PFV | T90 | -0.045 | -0.068 | -33. | FALSE | FALSE | TRUE | 1.3e-1 | 2.4e-4 | 3.1e-2 | 1.6e-5 |
| 42 | 3593 | 6217 | PFV | ArBdn | -0.096 | -0.21 | -53. | FALSE | FALSE | TRUE | 4.8e-1 | 6.6e-3 | 2.4e-1 | 9.8e-4 |
| 43 | 3591 | 6190 | PFV | ArThr | -0.23 | -0.46 | -50. | FALSE | FALSE | TRUE | 5.5e-1 | 4.8e-2 | 3.1e-1 | 1.3e-2 |
| 44 | 3588 | 6211 | PFV | HypBdn | -0.17 | -0.21 | -18. | FALSE | TRUE | TRUE | 2.3e-2 | 7.8e-10 | 2.1e-3 | 1.3e-11 |
| 45 | 3588 | 6199 | PFV | HRBdn | -0.17 | -0.22 | -24. | FALSE | TRUE | TRUE | 2.3e-2 | 7.3e-10 | 1.9e-3 | 8.8e-12 |
| 46 | 3593 | 6219 | PFV | VntBdn | -0.17 | -0.21 | -22. | FALSE | FALSE | TRUE | 6.6e-2 | 2.8e-8 | 1.0e-2 | 5.6e-10 |
| 47 | 3593 | 6214 | PFV | Vactive | 0.0022 | 0.032 | -93. | FALSE | FALSE | TRUE | 9.2e-1 | 4.3e-2 | 8.7e-1 | 1.0e-2 |
| 48 | 3594 | 6219 | PFV | Vpassive | 0.0080 | 0.055 | -85. | FALSE | FALSE | TRUE | 8.5e-1 | 3.3e-4 | 6.7e-1 | 2.4e-5 |
| 49 | 3593 | 6201 | OOO | LGN | -0.059 | -0.040 | 48. | FALSE | TRUE | TRUE | 1.3e-2 | 1.2e-2 | 6.0e-4 | 1.9e-3 |
| 50 | 3588 | 6211 | PBM | T90 | -0.064 | -0.080 | -21. | FALSE | TRUE | TRUE | 2.4e-2 | 1.8e-5 | 2.3e-3 | 6.7e-7 |
| 51 | 3588 | 6211 | PBM | HypBdn | -0.042 | -0.086 | -51. | FALSE | FALSE | TRUE | 6.7e-1 | 2.7e-2 | 4.4e-1 | 5.9e-3 |
| 52 | 3593 | 6219 | PBM | VntBdn | -0.073 | -0.090 | -19. | FALSE | FALSE | TRUE | 5.2e-1 | 4.5e-2 | 2.7e-1 | 1.2e-2 |
| 53 | 3588 | 6211 | VKL | T90 | 0.14 | 0.16 | -11. | FALSE | TRUE | TRUE | 1.1e-9 | 3.3e-23 | 4.6e-12 | 1.4e-25 |
| 54 | 3593 | 6201 | VKL | ChxDly | -0.39 | -0.58 | -34. | FALSE | FALSE | TRUE | 1.6e-1 | 5.5e-5 | 3.9e-2 | 3.1e-6 |
| 55 | 3588 | 6199 | VKL | HRBdn | -0.12 | -0.12 | 0.24 | FALSE | FALSE | TRUE | 1.1e-1 | 1.5e-3 | 2.4e-2 | 1.7e-4 |

|  | N<br>Sens. | N<br>Prim. | Topic<br>Abbr. | Exposure | Beta<br>Sens. | Beta<br>Prim. | %<br>Change | Sign<br>Change | FDR<br>< 0.05<br>Sens. | FDR<br>< 0.05<br>Prim. | FDR-adj.<br>P-value<br>Sens. | FDR-adj.<br>P-value<br>Prim. | P-value<br>Sens. | P-value<br>Prim. |
| --- | --- | --- | --- | --- | --- | --- | --- | --- | --- | --- | --- | --- | --- | --- |
| 56 | 3593 | 6201 | VKL | LG1 | 0.037 | 0.039 | -4.4 | FALSE | FALSE | TRUE | 1.7e-1 | 1.9e- 2 | 4.9e- 2 | 3.6e- 3 |
| 57 | 3593 | 6214 | VKL | Vactive | 0.057 | 0.061 | -7.1 | FALSE | TRUE | TRUE | 1.8e-3 | 1.8e- 5 | 5.7e- 5 | 7.5e- 7 |
| 58 | 3593 | 6214 | VKL | Vcomp | 0.064 | 0.061 | 4.4 | FALSE | TRUE | TRUE | 1.5e-4 | 1.8e- 5 | 3.0e- 6 | 7.0e- 7 |
| 59 | 3594 | 6219 | VKL | Vpassive | 0.019 | 0.035 | -44. | FALSE | FALSE | TRUE | 5.5e-1 | 3.1e- 2 | 3.1e- 1 | 7.0e- 3 |
| 60 | 3588 | 6211 | ILL | T90 | 0.10 | 0.12 | -14. | FALSE | TRUE | TRUE | 8.3e-5 | 5.9e-12 | 1.3e- 6 | 4.8e-14 |
| 61 | 3593 | 6201 | ILL | ChxDly | -0.42 | -0.40 | 5.3 | FALSE | FALSE | TRUE | 1.2e-1 | 1.1e- 2 | 2.7e- 2 | 1.8e- 3 |
| 62 | 3593 | 6201 | ILL | LGN | -0.054 | -0.058 | -5.9 | FALSE | TRUE | TRUE | 3.4e-2 | 3.8e- 4 | 4.2e- 3 | 2.9e- 5 |
| 63 | 3593 | 6214 | ILL | Vactive | 0.047 | 0.041 | 13. | FALSE | TRUE | TRUE | 2.0e-2 | 6.6e- 3 | 1.0e- 3 | 1.0e- 3 |
| 64 | 3593 | 6214 | ILL | Vcomp | 0.037 | 0.033 | 13. | FALSE | FALSE | TRUE | 5.2e-2 | 4.0e- 2 | 7.8e- 3 | 9.5e- 3 |
| 65 | 3594 | 6219 | ILL | Vpassive | 0.068 | 0.040 | 71. | FALSE | TRUE | TRUE | 1.0e-2 | 1.4e- 2 | 4.1e- 4 | 2.5e- 3 |
| 66 | 3593 | 6201 | SSS | ChxDly | 0.42 | 0.38 | 9.7 | FALSE | FALSE | TRUE | 1.2e-1 | 1.3e- 2 | 2.8e- 2 | 2.3e- 3 |

Selected sensitivity (Sens.) results for primary (Prim.) analysis associations with FDR < 0.05, relating ATM Topics (dependent variable) and PSG variables (exposures).

**Supplementary Table 4.** Sensitivity Analysis: Multi-exposure LASSO model.

|  | N<br>Sens. | N<br>Prim. | Topic<br>Abbr. | Exposure | Beta<br>Sens. | Beta<br>Prim. | %<br>Change | Sign<br>Change | FDR<br>< 0.05<br>Sens. | FDR<br>< 0.05<br>Prim. | FDR-adj.<br>P-value<br>Sens. | FDR-adj.<br>P-value<br>Prim. | P-value<br>Sens. | P-value<br>Prim. |
| --- | --- | --- | --- | --- | --- | --- | --- | --- | --- | --- | --- | --- | --- | --- |
| 1 | 6165 | 6199 | IPK | HRBdn | -0.021 | -0.091 | -77. | FALSE | FALSE | TRUE | 4.9e-1 | 2.4e-2 | 2.7e-1 | 4.6e-3 |
| 2 | 6165 | 6219 | IPK | VntBdn | -0.014 | -0.097 | -85. | FALSE | FALSE | TRUE | 7.0e-1 | 2.4e-2 | 4.6e-1 | 4.5e-3 |
| 3 | 6165 | 6214 | IPK | Vactive | — | 0.034 | — | — | — | TRUE | — | 2.6e-2 | — | 5.5e-3 |
| 4 | 6165 | 6219 | IPK | Vpassive | 0.0091 | 0.036 | -75. | FALSE | FALSE | TRUE | 7.5e-1 | 2.5e-2 | 5.2e-1 | 5.0e-3 |
| 5 | 6165 | 6211 | UMU | T90 | -0.038 | -0.052 | -28. | FALSE | FALSE | TRUE | 3.6e-1 | 6.6e-3 | 1.4e-1 | 1.0e-3 |
| 6 | 6165 | 6220 | IMG | AHI | — | 0.16 | — | — | — | TRUE | — | 8.2e-4 | — | 7.6e-5 |
| 7 | 6165 | 6190 | IMG | ArThr | — | 0.49 | — | — | — | TRUE | — | 2.6e-2 | — | 5.6e-3 |
| 8 | 6165 | 6211 | IMG | HypBdn | 0.10 | 0.13 | -22. | FALSE | TRUE | TRUE | 1.0e-2 | 8.8e-5 | 7.5e-4 | 5.4e-6 |
| 9 | 6165 | 6199 | IMG | HRBdn | 0.067 | 0.16 | -58. | FALSE | TRUE | TRUE | 1.6e-2 | 1.8e-5 | 1.5e-3 | 5.4e-7 |
| 10 | 6165 | 6219 | IMG | VntBdn | -0.085 | 0.13 | -167. | TRUE | TRUE | TRUE | 3.8e-2 | 1.4e-3 | 4.7e-3 | 1.4e-4 |
| 11 | 6165 | 6219 | IMG | Vpassive | -0.018 | -0.043 | -59. | FALSE | FALSE | TRUE | 5.2e-1 | 3.9e-3 | 2.8e-1 | 5.3e-4 |
| 12 | 6165 | 6211 | IVO | T90 | -0.051 | -0.051 | -0.66 | FALSE | TRUE | TRUE | 1.6e-2 | 3.9e-3 | 1.4e-3 | 5.1e-4 |
| 13 | 6165 | 6217 | IVO | ArBdn | -0.052 | -0.20 | -75. | FALSE | TRUE | TRUE | 3.7e-3 | 3.9e-3 | 1.6e-4 | 5.1e-4 |
| 14 | 6165 | 6211 | HVK | T90 | 0.064 | 0.075 | -15. | FALSE | TRUE | TRUE | 5.8e-3 | 5.1e-5 | 3.3e-4 | 2.7e-6 |
| 15 | 6165 | 6199 | HVK | HRBdn | -0.12 | -0.12 | -3.0 | FALSE | TRUE | TRUE | 6.3e-5 | 2.1e-3 | 1.6e-6 | 2.5e-4 |
| 16 | 6165 | 6201 | HVK | LGN | 0.048 | 0.035 | 37. | FALSE | FALSE | TRUE | 4.4e-1 | 4.5e-2 | 2.0e-1 | 1.1e-2 |
| 17 | 6165 | 6214 | HVK | Vactive | 0.039 | 0.042 | -5.5 | FALSE | FALSE | TRUE | 2.4e-1 | 6.6e-3 | 6.3e-2 | 1.0e-3 |
| 18 | 6165 | 6214 | HVK | Vcomp | 0.0028 | 0.034 | -92. | FALSE | FALSE | TRUE | 1.0e+0 | 3.1e-2 | 9.1e-1 | 7.1e-3 |
| 19 | 6165 | 6211 | BMV | T90 | 0.015 | 0.040 | -63. | FALSE | FALSE | TRUE | 5.5e-1 | 3.8e-2 | 3.3e-1 | 8.8e-3 |
| 20 | 6165 | 6214 | BMV | Vactive | 0.036 | 0.031 | 18. | FALSE | FALSE | TRUE | 2.3e-1 | 4.5e-2 | 5.8e-2 | 1.1e-2 |
| 21 | 6165 | 6214 | BMV | Vcomp | — | 0.031 | — | — | — | TRUE | — | 4.5e-2 | — | 1.1e-2 |
| 22 | 6165 | 6211 | DDD | T90 | -0.082 | -0.072 | 14. | FALSE | TRUE | TRUE | 2.2e-2 | 2.8e-5 | 2.2e-3 | 1.2e-6 |
| 23 | 6165 | 6220 | PPM | AHI | 0.14 | -0.10 | -229. | TRUE | FALSE | TRUE | 1 e+0 | 4.7e-2 | 1 e+0 | 1.2e-2 |
| 24 | 6165 | 6211 | PPM | HypBdn | -0.093 | -0.12 | -21. | FALSE | FALSE | TRUE | 1 e+0 | 9.3e-4 | 1 e+0 | 9.1e-5 |
| 25 | 6165 | 6199 | PPM | HRBdn | -0.054 | -0.13 | -58. | FALSE | FALSE | TRUE | 1 e+0 | 9.5e-4 | 1 e+0 | 9.6e-5 |
| 26 | 6165 | 6219 | PPM | VntBdn | -0.059 | -0.11 | -48. | FALSE | FALSE | TRUE | 1 e+0 | 6.6e-3 | 1 e+0 | 1.0e-3 |
| 27 | 6165 | 6214 | PPM | Vactive | -0.014 | 0.038 | -138. | TRUE | FALSE | TRUE | 1 e+0 | 1.2e-2 | 1 e+0 | 2.1e-3 |
| 28 | 6165 | 6214 | PPM | Vcomp | 0.035 | 0.035 | 1.3 | FALSE | FALSE | TRUE | 1 e+0 | 2.5e-2 | 1 e+0 | 5.1e-3 |
| 29 | 6165 | 6219 | PPM | Vpassive | 0.028 | 0.036 | -24. | FALSE | FALSE | TRUE | 1 e+0 | 2.6e-2 | 1 e+0 | 5.3e-3 |
| 30 | 6165 | 6220 | BMK | AHI | 0.0040 | 0.17 | -98. | FALSE | FALSE | TRUE | 9.1e-1 | 1.5e-4 | 7.7e-1 | 9.6e-6 |
| 31 | 6165 | 6190 | BMK | ArThr | 0.0073 | 0.61 | -99. | FALSE | FALSE | TRUE | 7.3e-1 | 2.3e-3 | 4.9e-1 | 2.8e-4 |
| 32 | 6165 | 6201 | BMK | ChxDly | -0.030 | -0.44 | -93. | FALSE | FALSE | TRUE | 1.3e-1 | 1.4e-3 | 2.0e-2 | 1.5e-4 |
| 33 | 6165 | 6211 | BMK | HypBdn | 0.056 | 0.13 | -58. | FALSE | FALSE | TRUE | 1.2e-1 | 4.2e-5 | 1.8e-2 | 2.0e-6 |
| 34 | 6165 | 6199 | BMK | HRBdn | -0.0099 | 0.090 | -111. | TRUE | FALSE | TRUE | 8.1e-1 | 1.5e-2 | 6.2e-1 | 2.8e-3 |
| 35 | 6165 | 6201 | BMK | LG1 | 0.016 | 0.050 | -69. | FALSE | FALSE | TRUE | 4.9e-1 | 4.4e-4 | 2.6e-1 | 3.6e-5 |
| 36 | 6165 | 6219 | BMK | VntBdn | — | 0.13 | — | — | — | TRUE | — | 8.1e-4 | — | 7.2e-5 |
| 37 | 6165 | 6219 | BMK | Vpassive | -0.003 | -0.037 | -92. | FALSE | FALSE | TRUE | 8.7e-1 | 1.0e-2 | 7.0e-1 | 1.6e-3 |
| 38 | 6165 | 6201 | IIL | LG1 | -0.032 | -0.033 | -4.2 | FALSE | FALSE | TRUE | 5.1e-2 | 3.8e-2 | 6.7e-3 | 9.0e-3 |
| 39 | 6165 | 6211 | IIC | T90 | -0.081 | -0.062 | 32. | FALSE | TRUE | TRUE | 1.1e-5 | 6.7e-4 | 2.1e-7 | 5.7e-5 |
| 40 | 6165 | 6220 | PFV | AHI | — | -0.25 | — | — | — | TRUE | — | 7.2e-8 | — | 1.8e-9 |
| 41 | 6165 | 6211 | PFV | T90 | -0.027 | -0.068 | -61. | FALSE | FALSE | TRUE | 5.4e-1 | 2.4e-4 | 3.2e-1 | 1.6e-5 |
| 42 | 6165 | 6217 | PFV | ArBdn | -0.0066 | -0.21 | -97. | FALSE | FALSE | TRUE | 8.1e-1 | 6.6e-3 | 6.2e-1 | 9.8e-4 |
| 43 | 6165 | 6190 | PFV | ArThr | 0.014 | -0.46 | -103. | TRUE | FALSE | TRUE | 4.8e-1 | 4.8e-2 | 2.5e-1 | 1.3e-2 |
| 44 | 6165 | 6211 | PFV | HypBdn | -0.045 | -0.21 | -78. | FALSE | FALSE | TRUE | 2.7e-1 | 7.8e-10 | 8.4e-2 | 1.3e-11 |
| 45 | 6165 | 6199 | PFV | HRBdn | -0.052 | -0.22 | -77. | FALSE | FALSE | TRUE | 7.3e-2 | 7.3e-10 | 1.0e-2 | 8.8e-12 |
| 46 | 6165 | 6219 | PFV | VntBdn | — | -0.21 | — | — | — | TRUE | — | 2.8e-8 | — | 5.6e-10 |
| 47 | 6165 | 6214 | PFV | Vactive | — | 0.032 | — | — | — | TRUE | — | 4.3e-2 | — | 1.0e-2 |
| 48 | 6165 | 6219 | PFV | Vpassive | — | 0.055 | — | — | — | TRUE | — | 3.3e-4 | — | 2.4e-5 |
| 49 | 6165 | 6201 | OOO | LGN | -0.042 | -0.040 | 5.9 | FALSE | FALSE | TRUE | 4.8e-1 | 1.2e-2 | 2.5e-1 | 1.9e-3 |
| 50 | 6165 | 6211 | PBM | T90 | -0.052 | -0.080 | -35. | FALSE | TRUE | TRUE | 2.2e-4 | 1.8e-5 | 8.2e-6 | 6.7e-7 |
| 51 | 6165 | 6211 | PBM | HypBdn | — | -0.086 | — | — | — | TRUE | — | 2.7e-2 | — | 5.9e-3 |
| 52 | 6165 | 6219 | PBM | VntBdn | — | -0.090 | — | — | — | TRUE | — | 4.5e-2 | — | 1.2e-2 |
| 53 | 6165 | 6211 | VKL | T90 | 0.14 | 0.16 | -12. | FALSE | FALSE | TRUE | 2.8e-1 | 3.3e-23 | 8.9e-2 | 1.4e-25 |
| 54 | 6165 | 6201 | VKL | ChxDly | -0.049 | -0.58 | -92. | FALSE | TRUE | TRUE | 1.0e-2 | 5.5e-5 | 7.1e-4 | 3.1e-6 |
| 55 | 6165 | 6199 | VKL | HRBdn | -0.14 | -0.12 | 15. | FALSE | TRUE | TRUE | 5.0e-6 | 1.5e-3 | 6.2e-8 | 1.7e-4 |
| 56 | 6165 | 6201 | VKL | LG1 | 0.021 | 0.039 | -45. | FALSE | FALSE | TRUE | 3.6e- 1 | 1.9e- 2 | 1.4e- 1 | 3.6e- 3 |
| 57 | 6165 | 6214 | VKL | Vactive | — | 0.061 | — | — | — | TRUE | — | 1.8e- 5 | — | 7.5e- 7 |
| 58 | 6165 | 6214 | VKL | Vcomp | 0.058 | 0.061 | -5.7 | FALSE | TRUE | TRUE | 1.6e- 4 | 1.8e- 5 | 4.9e- 6 | 7.0e- 7 |
| 59 | 6165 | 6219 | VKL | Vpassive | 0.023 | 0.035 | -34. | FALSE | FALSE | TRUE | 3.3e- 1 | 3.1e- 2 | 1.1e- 1 | 7.0e- 3 |
| 60 | 6165 | 6211 | ILL | T90 | 0.10 | 0.12 | -13. | FALSE | TRUE | TRUE | 6.2e-10 | 5.9e-12 | 3.9e-12 | 4.8e-14 |
| 61 | 6165 | 6201 | ILL | ChxDly | -0.027 | -0.40 | -93. | FALSE | FALSE | TRUE | 2.0e- 1 | 1.1e- 2 | 4.0e- 2 | 1.8e- 3 |
| 62 | 6165 | 6201 | ILL | LGN | -0.051 | -0.058 | -11. | FALSE | FALSE | TRUE | 1.8e- 1 | 3.8e- 4 | 3.1e- 2 | 2.9e- 5 |
| 63 | 6165 | 6214 | ILL | Vactive | 0.015 | 0.041 | -63. | FALSE | FALSE | TRUE | 8.1e- 1 | 6.6e- 3 | 6.1e- 1 | 1.0e- 3 |
| 64 | 6165 | 6214 | ILL | Vcomp | 0.020 | 0.033 | -37. | FALSE | FALSE | TRUE | 5.2e- 1 | 4.0e- 2 | 2.9e- 1 | 9.5e- 3 |
| 65 | 6165 | 6219 | ILL | Vpassive | 0.032 | 0.040 | -20. | FALSE | FALSE | TRUE | 3.6e- 1 | 1.4e- 2 | 1.4e- 1 | 2.5e- 3 |
| 66 | 6165 | 6201 | SSS | ChxDly | 0.037 | 0.38 | -90. | FALSE | FALSE | TRUE | 5.7e- 2 | 1.3e- 2 | 7.9e- 3 | 2.3e- 3 |

Selected sensitivity results for primary analysis associations with FDR < 0.05, relating ATM Topics (dependent variable) and PSG variables (exposures).

